# Condom use and HIV testing among adults in Switzerland: repeated national cross-sectional surveys 2007, 2012, and 2017

**DOI:** 10.1101/2022.12.05.22283096

**Authors:** Diana Buitrago-Garcia, Georgia Salanti, Nicola Low

**Affiliations:** Institute of Social and Preventive Medicine, University of Bern, Bern, Switzerland

**Keywords:** Condoms, HIV, prevalence, health surveys

## Abstract

**Background:** Monitoring of HIV and sexually transmitted infection (STI) prevention is important for guiding national sexual health programmes for both the general population and key populations. The objectives of this study were to examine patterns of condom use at last intercourse and lifetime HIV testing from 2007 to 2017 in Switzerland, and to explore factors associated with these behaviours in men and women with opposite-sex partners and with same sex partners.

**Methods:** We analysed data from the 2007, 2012 and 2017 Swiss Health Survey. At each time point, outcome and population group, we conducted a descriptive analysis of weighted data and conducted multivariable logistic regression to obtain adjusted odds ratios (aOR) with 95% confidence intervals (CI) and compared outcomes between the timepoints.

**Results:** In total, 43,949 people were interviewed: 21,274 men and 22,675 women, who reported having sex only with partners of the opposite sex, 633 men who reported sex with a male partner and 699 women who reported sex with a female partner. Among the three surveys the prevalence of condom use varied from 24 to 26% of men and 18 to 21% in women with only opposite-sex partners (aOR men, 0.93, 95% CI 0.82, 1.06; women 0.98, 95% CI 0.86 to 1.11). In men with any same sex partner the prevalence of condom use increased from 43% in 2007 to 54% in 2017 (aOR 1.80, 95% CI 0.97, 3.34). In multivariable analysis, the factor most strongly associated with condom use was sex with an occasional partner at last intercourse. HIV testing ever increased across all three survey years in all groups: 2017 vs. 2007, aOR men with only opposite-sex partners 1.57 (95% CI 1.42, 1.74), women with only opposite-sex partners 1.54 (1.39, 1.71), men with any same sex partner 1.85 (0.96, 3.55), women with any same sex partner 1.31 (0.74, 2.30).

**Conclusions:** Monitoring of condom use, and HIV testing should continue and contribute to the development of the national sexual health programme. Stronger promotion of condoms for people with opposite-sex partners might be needed, since overall condom use at last intercourse has not changed since 2007.

## Background

Monitoring of HIV and sexually transmitted infection (STI) prevention is important for guiding national sexual health strategies for both the general population and key populations,(1) including men who have sex with men, who are disproportionately affected by HIV and other STIs.(2) Consistent condom use is an important and effective method for the prevention of many STIs, including HIV, although their effectiveness varies according to the infection.(3, 4) Testing for antibodies to HIV infection allows early identification of new HIV infections, which enables earlier access to antiretroviral therapy. A negative test for HIV can now allow access to pre-exposure prophylactic (PrEP) medication for people whose sexual lifestyle or practices increase the risk of acquiring HIV, and whose use has been increasing since 2015.(5, 6)

Surveys of nationally representative samples contribute to understanding changing patterns of sexual behaviours and preventive practices at the population level.(7) In the 1990 and 2000 British National Surveys of Sexual Attitudes and Lifestyles (Natsal), condom use in the past year reported by 16 to 44 year olds increased from 43 to 51% in men and from 31 to 39% in women.(8) In the United States of America (USA), around 20% of men and women aged 18 years and older reported condom use at last sexual intercourse with little change across biennial General Social Surveys from 1996 to 2008.(9) Men who have sex with men report higher levels of condom use and HIV testing than men who have only opposite-sex partners.(10, 11) Few studies examine patterns of condom use and HIV testing in women with female partners as HIV risk is lower in this group and condom use is thought to be less relevant.(12)

Condom use and HIV testing have been monitored as key indicators in the national strategy for the prevention of HIV and STIs in Switzerland since 1987.(13) The Swiss Federal Office of Public Health incorporated evaluations into a system of behavioural surveillance, as recommended for countries with concentrated epidemics of HIV, in 2004.(14) The objectives of this study were to examine patterns of condom use and HIV testing from 2007 to 2017 in Switzerland and to explore factors associated with these preventive behaviours in groups of people with opposite-sex partners and with same sex partners, using data from population-based surveys.

## Methods

### Study design

We analysed all three rounds of the Swiss Health Survey that included questions on condom use and HIV testing since 2007 when national monitoring of HIV and STI prevention was incorporated into the five-yearly Swiss Health Survey.(15) The Swiss Health Survey is a cross-sectional survey conducted every five years since 1992, with a representative sample of the permanent resident adult population aged 15 years and older who speak German, French, or Italian.(15) People living in an institution (hospital, nursing home, prison, convent or monastery) or resident in Switzerland for 3 months or fewer at the time of the survey were not invited. The Swiss Federal Statistical Office used the sampling frame for personal and household surveys, which is based on data from cantonal and municipal registers of residents and supplemented quarterly with information from telephone service providers. The sample is generated for each survey through stratified, multistage random sampling in each canton. A sample weight is then assigned to each observation, according to region, household size, age, sex and nationality.

### Study groups and variables

We considered respondents in four sexual behaviour groups, using information available in all three surveys. We stratified respondents by sex (men and women) and by the reported sex of their sexual partners: men with only female partners, women with only male partners, men with any male partner, and women with any female partner. The questions about sexual partners referred to the sexual lifetime for the surveys in 2007 and 2012 and the last five years for the 2017 survey. There was no category for people with a non-binary gender identity. A separate question about sexual orientation was asked for the first time in 2017.

#### Outcome variables

We examined two outcomes, condom use at last sexual intercourse and ever having had an HIV test. The questions were asked by an interviewer by telephone to participants between 16 and 74 years old and additional filtering questions determined the group of eligible respondents for each question (Figure 1). Some filtering questions differed between surveys (Table S1). For condom use, the question asked, “Did you use a male condom the last time you had sex?” For testing for HIV, the question asked, “How many times have you taken an AIDS test? and “When was the last time?”

**Figure 1.**
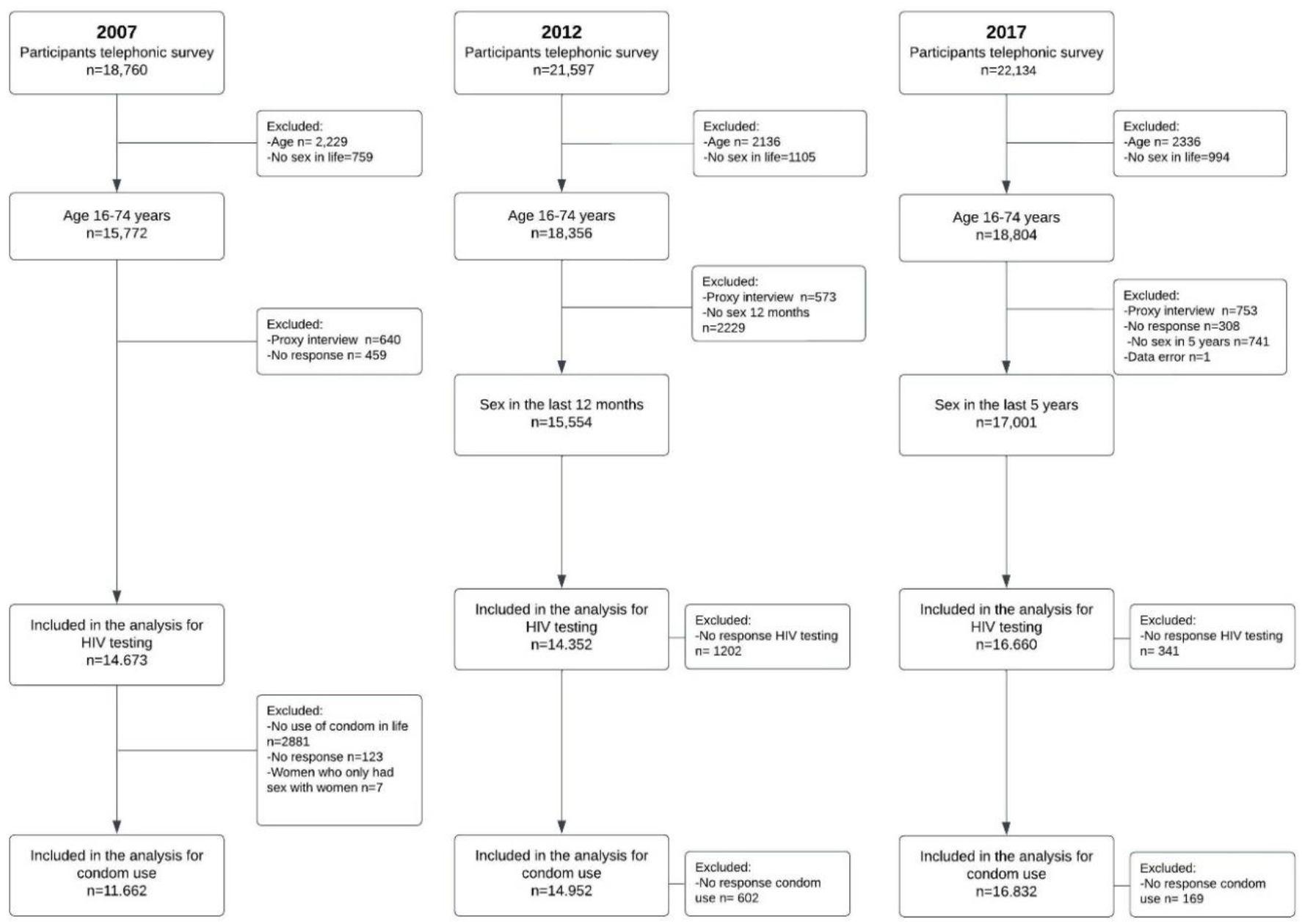
Flowchart of inclusion and exclusions for respondents to questions about condom use and HIV testing in the Swiss Health Survey 2007, 2012 and 2017

#### Exposure variables

We selected variables from the questionnaire, which we considered *a priori* to be relevant to condom use and HIV testing, in the following categories: demographic (age), social and economic (region of residence, education level, personal income and marital status); sexual behaviours (age at first sexual intercourse, frequency of sexual intercourse during the last 12 months, number of partners during the last 12 months and if the partner was stable, occasional or a sex worker); and other behaviours (consumption of alcohol, tobacco, cannabis, and other illicit drugs, heroin, cocaine, or ecstasy).

### Statistical analysis

We used the *survey*(16) package in R (version 3.5.1) to incorporate the weightings provided by the Swiss Federal Statistical Office(17) and to generate results that are representative of the Swiss population. For each survey and outcome, we conducted a descriptive analysis of the weighted data and reported percentages (with 95% confidence intervals) for each categorical exposure variable, the median and interquartile range (IQR) for continuous variables and unadjusted odds ratios (OR, 95% CI) for associations between each outcome and exposures.

We then conducted multivariable logistic regression analyses to obtain adjusted odds ratios (aOR, 95% CI) for the associations between each outcome and exposure variable. We added survey year as a variable to examine evidence of a change over time. In the multivariable analyses of condom use, we examine the change only between the 2012 and 2017 surveys because in 2007 there was an additional exclusion criterion. People who had never used a condom were not asked about the use of condoms at last sexual intercourse (Figure 1). For men and women with only opposite-sex partners, the multivariable model includes all exposure variables and the year of the survey. For people with same sex partners, the regression model included only survey year, age, number of sex partners and type of partner in the multivariable model, owing to the small sample sizes.

For each outcome, we report findings separately for each of the four sexual behaviour groups. For analyses of HIV testing, we include all three survey years in multivariable analyses because eligibility criteria for responding to the question were similar.

## Results

In the three surveys, a total of 43,949 people were interviewed by telephone, comprising 21,274 men and 22,675 women who reported having only sex partners of the opposite sex, 633 men who reported any male sex partner and 699 women who reported any female sex partner (Figure 1, Table S2).

Table 1 summarises the overall unweighted and weighted denominators and prevalence for each outcome and each study group. The proportion of respondents reporting condom use at last sexual intercourse appeared similar in each survey year: men (24 to 26%) and women (18 to 21%) with only opposite-sex partners; men reporting any same sex partner (33 to 54%), women reporting any same sex partner (most of whom had had sex with at least one man, 23 to 27%). Proportions of respondents reporting ever having had an HIV test appeared to increase over time: men (32 to 45%) and women (35 to 51%) with only opposite-sex partners; men (72 to 83%) and women (61 to 70%) reporting any same sex partner.

**Table 1.** Prevalence of condom use at last sexual intercourse and ever having had an HIV test in Switzerland in 2007, 2012 and 2017

| Survey year | 2007 |  | 2012 |  | 2017 |  |
| --- | --- | --- | --- | --- | --- | --- |
|  | Denominator<br>N unweighted,<br>weighted | Prevalence<br>% (95% CI) | Denominator<br>N unweighted,<br>weighted | Prevalence<br>% (95% CI) | Denominator<br>N unweighted,<br>weighted | Prevalence<br>% (95% CI) |
| **Condom use at last sexual intercourse<sup>a</sup>** |  |  |  |  |  |  |
| Men, only female partners | 5415, 1972621 | 26 (24-27) | 7382, 2427588 | 24 (22-25) | 8049, 2724669 | 26 (25-28) |
| Women, only male partners | 5851, 1830947 | 20 (18-21) | 7025, 2132003 | 18 (17-19) | 8428, 2557958 | 21 (20-22) |
| Men, any male partner | 191, 66118 | 43 (34-53) | 245, 84238 | 33 (26-41) | 183, 76438 | 54 (46-63) |
| Women, any female partner<sup>b</sup> | 205, 63179 | 23 (17-31) | 300, 96257 | 27 (21-33) | 172, 69018 | 25 (18-33) |
| **HIV testing ever<sup>a</sup>** |  |  |  |  |  |  |
| Men, only female partners | 6236, 2237900 | 32 (30-33) | 7063, 2313010 | 42 (40-43) | 7975, 2702130 | 45 (44-46) |
| Women, only male partners | 7582, 2289504 | 35 (34-37) | 6763, 2049462 | 46 (45-48) | 8330, 2529129 | 51 (50-53) |
| Men, any male partner | 187, 64684 | 72 (64-80) | 237, 81464 | 80 (74-86) | 178, 74070 | 83 (77-89) |
| Women, any female partner | 216, 67259 | 61 (53-69) | 289, 92734 | 70 (63-77) | 172, 69018 | 69 (60-77) |

a. Overall denominators for condom use and HIV testing differ because eligibility criteria for answering each question differed;
b. In 2007, women who reported only female sex partners were not asked about condom use (n=7)
Abbreviations: CI, confidence interval.

### Condom use at last sexual intercourse

#### Men and women reporting only opposite-sex partners

Table 2 shows reported condom use at the last sexual intercourse and the aORs from the multivariable regression model comparing 2012 and 2017 (all results for 2007, Table S3, S4, unadjusted ORs 2012 and 2017, Table S3, S5). In all survey years, condom use was highest in the age group 16-24 years for both for men (from 60 to 67% in the three survey years) and women (43 to 48%), decreasing with increasing age, and higher among single (46 to 48% of men and 36 to 38% of women) than married, widowed, or divorced people, with little regional variability (Table 2, Table S4). Among men, condom use was reported most frequently by those who did not attend school or who only completed primary school and by those in the lowest income category (50 to 58% of those with no monthly income). Among women, reported condom use was similar according to education and income in all years. Condom use at last sexual intercourse was consistently higher in both women and men who reported higher numbers of sexual partners across all survey years. Among those reporting 5 or more partners in the last 12 months, 65 to 73% of men and 40 to 61% of women reported condom use, compared with <20% in people reporting 1 partner in the last 12 months. In 2012 and 2017 (question not asked in 2007), those reporting an occasional partner reported higher levels of condom use with that partner (68 to 81%) than those with a stable partner (≤ 20%). The pattern of condom use according to age at first sexual intercourse was not consistent across survey years and did not vary substantially according to alcohol and tobacco consumption but was higher in current than non-current or never users of cannabis and other illicit drugs. People who had ever tested for HIV were slightly more likely than those who had never tested for HIV to report using a condom.

**Table 2.**
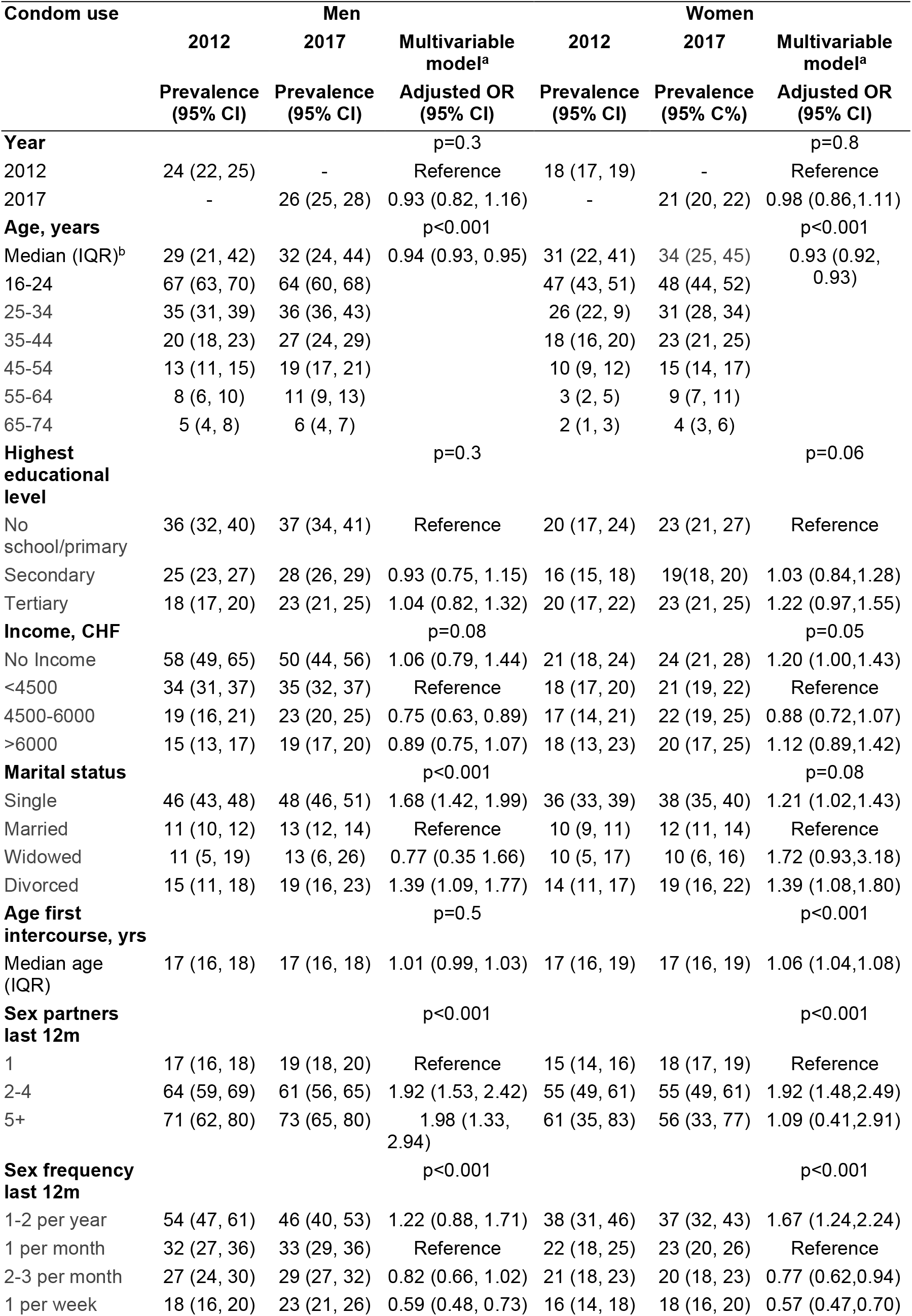
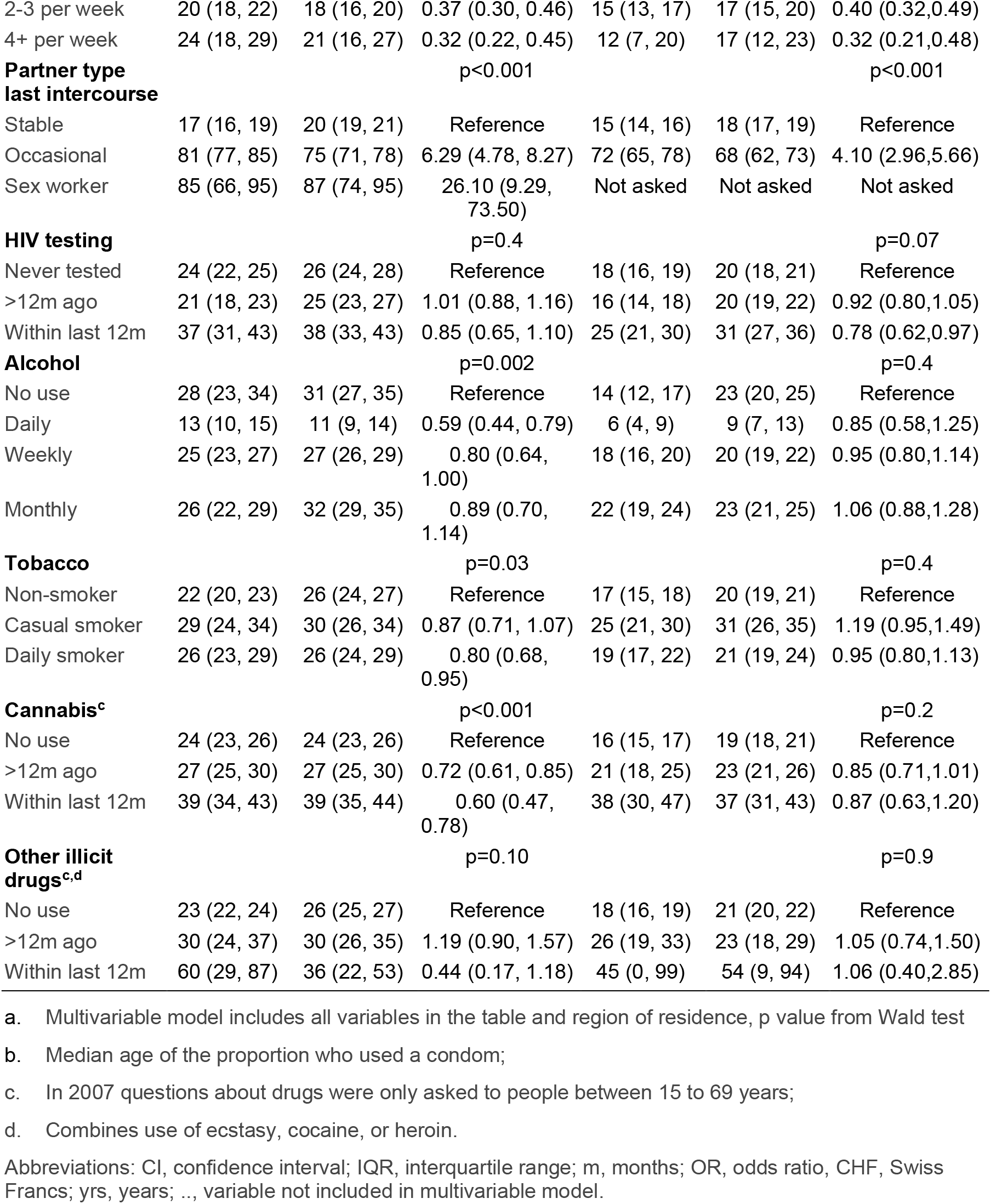
Prevalence of, and factors associated with, condom use at last intercourse, by sociodemographic and behavioural characteristics in men and women reporting only opposite-sex partners 2012 and 2017

| Condom use | Men |  |  | Women |  |  |
| --- | --- | --- | --- | --- | --- | --- |
|  | 2012 | 2017 | Multivariable model<sup>a</sup> | 2012 | 2017 | Multivariable model<sup>a</sup> |
|  | Prevalence (95% CI) | Prevalence (95% CI) | Adjusted OR (95% CI) | Prevalence (95% CI) | Prevalence (95% CI) | Adjusted OR (95% CI) |
| **Year** |  |  | p=0.3 |  |  | p=0.8 |
| 2012 | 24 (22, 25) | - | Reference | 18 (17, 19) | - | Reference |
| 2017 | - | 26 (25, 28) | 0.93 (0.82, 1.16) | - | 21 (20, 22) | 0.98 (0.86,1.11) |
| **Age, years** |  |  | p<0.001 |  |  | p<0.001 |
| Median (IQR)<sup>b</sup> | 29 (21, 42) | 32 (24, 44) | 0.94 (0.93, 0.95) | 31 (22, 41) | 34 (25, 45) | 0.93 (0.92, 0.93) |
| 16-24 | 67 (63, 70) | 64 (60, 68) |  | 47 (43, 51) | 48 (44, 52) |  |
| 25-34 | 35 (31, 39) | 36 (36, 43) |  | 26 (22, 9) | 31 (28, 34) |  |
| 35-44 | 20 (18, 23) | 27 (24, 29) |  | 18 (16, 20) | 23 (21, 25) |  |
| 45-54 | 13 (11, 15) | 19 (17, 21) |  | 10 (9, 12) | 15 (14, 17) |  |
| 55-64 | 8 (6, 10) | 11 (9, 13) |  | 3 (2, 5) | 9 (7, 11) |  |
| 65-74 | 5 (4, 8) | 6 (4, 7) |  | 2 (1, 3) | 4 (3, 6) |  |
| **Highest educational level** |  |  | p=0.3 |  |  | p=0.06 |
| No school/primary | 36 (32, 40) | 37 (34, 41) | Reference | 20 (17, 24) | 23 (21, 27) | Reference |
| Secondary | 25 (23, 27) | 28 (26, 29) | 0.93 (0.75, 1.15) | 16 (15, 18) | 19(18, 20) | 1.03 (0.84,1.28) |
| Tertiary | 18 (17, 20) | 23 (21, 25) | 1.04 (0.82, 1.32) | 20 (17, 22) | 23 (21, 25) | 1.22 (0.97,1.55) |
| **Income, CHF** |  |  | p=0.08 |  |  | p=0.05 |
| No Income | 58 (49, 65) | 50 (44, 56) | 1.06 (0.79, 1.44) | 21 (18, 24) | 24 (21, 28) | 1.20 (1.00,1.43) |
| <4500 | 34 (31, 37) | 35 (32, 37) | Reference | 18 (17, 20) | 21 (19, 22) | Reference |
| 4500-6000 | 19 (16, 21) | 23 (20, 25) | 0.75 (0.63, 0.89) | 17 (14, 21) | 22 (19, 25) | 0.88 (0.72,1.07) |
| >6000 | 15 (13, 17) | 19 (17, 20) | 0.89 (0.75, 1.07) | 18 (13, 23) | 20 (17, 25) | 1.12 (0.89,1.42) |
| **Marital status** |  |  | p<0.001 |  |  | p=0.08 |
| Single | 46 (43, 48) | 48 (46, 51) | 1.68 (1.42, 1.99) | 36 (33, 39) | 38 (35, 40) | 1.21 (1.02,1.43) |
| Married | 11 (10, 12) | 13 (12, 14) | Reference | 10 (9, 11) | 12 (11, 14) | Reference |
| Widowed | 11 (5, 19) | 13 (6, 26) | 0.77 (0.35 1.66) | 10 (5, 17) | 10 (6, 16) | 1.72 (0.93,3.18) |
| Divorced | 15 (11, 18) | 19 (16, 23) | 1.39 (1.09, 1.77) | 14 (11, 17) | 19 (16, 22) | 1.39 (1.08,1.80) |
| **Age first intercourse, yrs** |  |  | p=0.5 |  |  | p<0.001 |
| Median age (IQR) | 17 (16, 18) | 17 (16, 18) | 1.01 (0.99, 1.03) | 17 (16, 19) | 17 (16, 19) | 1.06 (1.04,1.08) |
| **Sex partners last 12m** |  |  | p<0.001 |  |  | p<0.001 |
| 1 | 17 (16, 18) | 19 (18, 20) | Reference | 15 (14, 16) | 18 (17, 19) | Reference |
| 2-4 | 64 (59, 69) | 61 (56, 65) | 1.92 (1.53, 2.42) | 55 (49, 61) | 55 (49, 61) | 1.92 (1.48,2.49) |
| 5+ | 71 (62, 80) | 73 (65, 80) | 1.98 (1.33, 2.94) | 61 (35, 83) | 56 (33, 77) | 1.09 (0.41,2.91) |
| **Sex frequency last 12m** |  |  | p<0.001 |  |  | p<0.001 |
| 1-2 per year | 54 (47, 61) | 46 (40, 53) | 1.22 (0.88, 1.71) | 38 (31, 46) | 37 (32, 43) | 1.67 (1.24,2.24) |
| 1 per month | 32 (27, 36) | 33 (29, 36) | Reference | 22 (18, 25) | 23 (20, 26) | Reference |
| 2-3 per month | 27 (24, 30) | 29 (27, 32) | 0.82 (0.66, 1.02) | 21 (18, 23) | 20 (18, 23) | 0.77 (0.62,0.94) |
| 1 per week | 18 (16, 20) | 23 (21, 26) | 0.59 (0.48, 0.73) | 16 (14, 18) | 18 (16, 20) | 0.57 (0.47,0.70) |
| 2-3 per week | 20 (18, 22) | 18 (16, 20) | 0.37 (0.30, 0.46) | 15 (13, 17) | 17 (15, 20) | 0.40 (0.32,0.49) |
| 4+ per week | 24 (18, 29) | 21 (16, 27) | 0.32 (0.22, 0.45) | 12 (7, 20) | 17 (12, 23) | 0.32 (0.21,0.48) |
| **Partner type last intercourse** |  |  | p<0.001 |  |  | p<0.001 |
| Stable | 17 (16, 19) | 20 (19, 21) | Reference | 15 (14, 16) | 18 (17, 19) | Reference |
| Occasional | 81 (77, 85) | 75 (71, 78) | 6.29 (4.78, 8.27) | 72 (65, 78) | 68 (62, 73) | 4.10 (2.96,5.66) |
| Sex worker | 85 (66, 95) | 87 (74, 95) | 26.10 (9.29, 73.50) | Not asked | Not asked | Not asked |
| **HIV testing** |  |  | p=0.4 |  |  | p=0.07 |
| Never tested | 24 (22, 25) | 26 (24, 28) | Reference | 18 (16, 19) | 20 (18, 21) | Reference |
| >12m ago | 21 (18, 23) | 25 (23, 27) | 1.01 (0.88, 1.16) | 16 (14, 18) | 20 (19, 22) | 0.92 (0.80,1.05) |
| Within last 12m | 37 (31, 43) | 38 (33, 43) | 0.85 (0.65, 1.10) | 25 (21, 30) | 31 (27, 36) | 0.78 (0.62,0.97) |
| **Alcohol** |  |  | p=0.002 |  |  | p=0.4 |
| No use | 28 (23, 34) | 31 (27, 35) | Reference | 14 (12, 17) | 23 (20, 25) | Reference |
| Daily | 13 (10, 15) | 11 (9, 14) | 0.59 (0.44, 0.79) | 6 (4, 9) | 9 (7, 13) | 0.85 (0.58,1.25) |
| Weekly | 25 (23, 27) | 27 (26, 29) | 0.80 (0.64, 1.00) | 18 (16, 20) | 20 (19, 22) | 0.95 (0.80,1.14) |
| Monthly | 26 (22, 29) | 32 (29, 35) | 0.89 (0.70, 1.14) | 22 (19, 24) | 23 (21, 25) | 1.06 (0.88,1.28) |
| **Tobacco** |  |  | p=0.03 |  |  | p=0.4 |
| Non-smoker | 22 (20, 23) | 26 (24, 27) | Reference | 17 (15, 18) | 20 (19, 21) | Reference |
| Casual smoker | 29 (24, 34) | 30 (26, 34) | 0.87 (0.71, 1.07) | 25 (21, 30) | 31 (26, 35) | 1.19 (0.95,1.49) |
| Daily smoker | 26 (23, 29) | 26 (24, 29) | 0.80 (0.68, 0.95) | 19 (17, 22) | 21 (19, 24) | 0.95 (0.80,1.13) |
| **Cannabis<sup>c</sup>** |  |  | p<0.001 |  |  | p=0.2 |
| No use | 24 (23, 26) | 24 (23, 26) | Reference | 16 (15, 17) | 19 (18, 21) | Reference |
| >12m ago | 27 (25, 30) | 27 (25, 30) | 0.72 (0.61, 0.85) | 21 (18, 25) | 23 (21, 26) | 0.85 (0.71,1.01) |
| Within last 12m | 39 (34, 43) | 39 (35, 44) | 0.60 (0.47, 0.78) | 38 (30, 47) | 37 (31, 43) | 0.87 (0.63,1.20) |
| **Other illicit drugs<sup>c,d</sup>** |  |  | p=0.10 |  |  | p=0.9 |
| No use | 23 (22, 24) | 26 (25, 27) | Reference | 18 (16, 19) | 21 (20, 22) | Reference |
| >12m ago | 30 (24, 37) | 30 (26, 35) | 1.19 (0.90, 1.57) | 26 (19, 33) | 23 (18, 29) | 1.05 (0.74,1.50) |
| Within last 12m | 60 (29, 87) | 36 (22, 53) | 0.44 (0.17, 1.18) | 45 (0, 99) | 54 (9, 94) | 1.06 (0.40,2.85) |

a. Multivariable model includes all variables in the table and region of residence, p value from Wald test
b. Median age of the proportion who used a condom;
c. In 2007 questions about drugs were only asked to people between 15 to 69 years;
d. Combines use of ecstasy, cocaine, or heroin.
Abbreviations: CI, confidence interval; IQR, interquartile range; m, months; OR, odds ratio, CHF, Swiss Francs; yrs, years; ..., variable not included in multivariable model.

Comparing responses in 2017 with 2012 in multivariable analysis, there was no change in reported condom use at last sexual intercourse in either men (aOR 0.93, 95% CI 0.82 to 1.06) or women (aOR 0.98, 95% CI 0.86 to 1.11). The strength of associations with most exposure variables was attenuated (Table S5) except for age (for each year of increase in men, aOR 0.94, 95% CI 0.93, 0.95 and women, aOR 0.93, 95% CI 0.92, 0.93) and sex with an occasional rather than a stable partner at last sexual intercourse (aOR men 6.29, 95% CI 4.78, 8.27; aOR women 4.10, 95% CI 2.96, 5.66) remained most strongly associated with condom use. Amongst men whose last sexual partner was a sex worker, the adjusted odds of condom use were 26.1 (95% CI 9.29, 73.5) times higher than for use with a stable partner.

#### Men reporting any same sex partner

Table 3 shows the prevalence of reported condom use at last sexual intercourse for all survey years. In all survey years, condom use at last sexual intercourse was >80% in 16 to 24 year olds, decreasing with the age of the respondent. The pattern of condom use according to other sociodemographic factors was similar to that in men reporting only opposite-sex partners. Among men reporting 5 or more partners in the last 12 months, 64% in 2007, 84% in 2012 and 86% in 2017 reported using a condom at last sexual intercourse, compared with 18 to 31% of those reporting 1 partner. In 2012 and 2017, reported condom use was higher with an occasional (74% and 83%, respectively) than a stable partner (23% and 33%). Reported condom use at last sex was most common amongst men who had had a test for HIV in the 12 months before the survey, 61% in 2007, 58% in 2012 and 63% in 2017, and amongst those reporting current alcohol, tobacco, cannabis or other illicit drug use (Table S6).

**Table 3.**
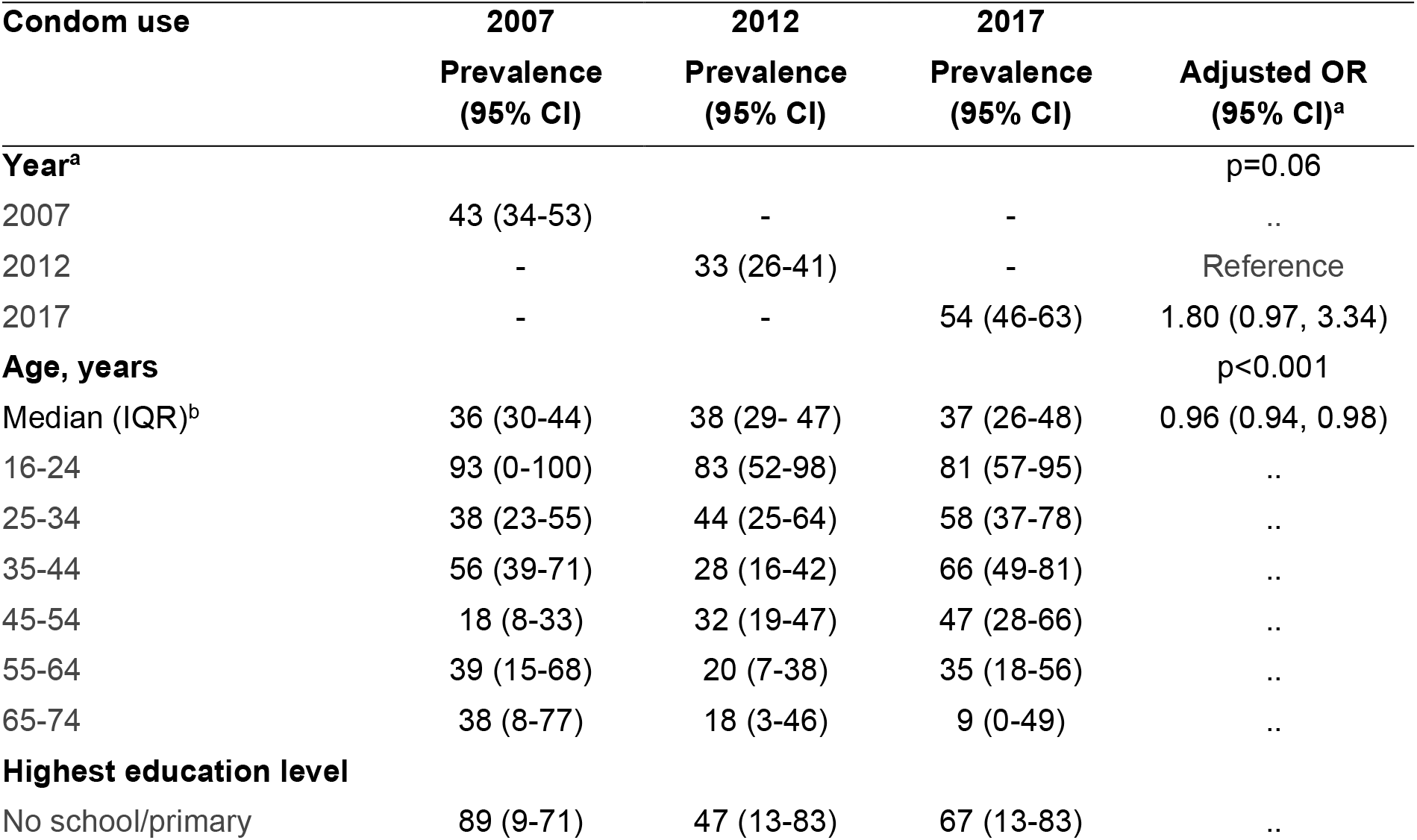
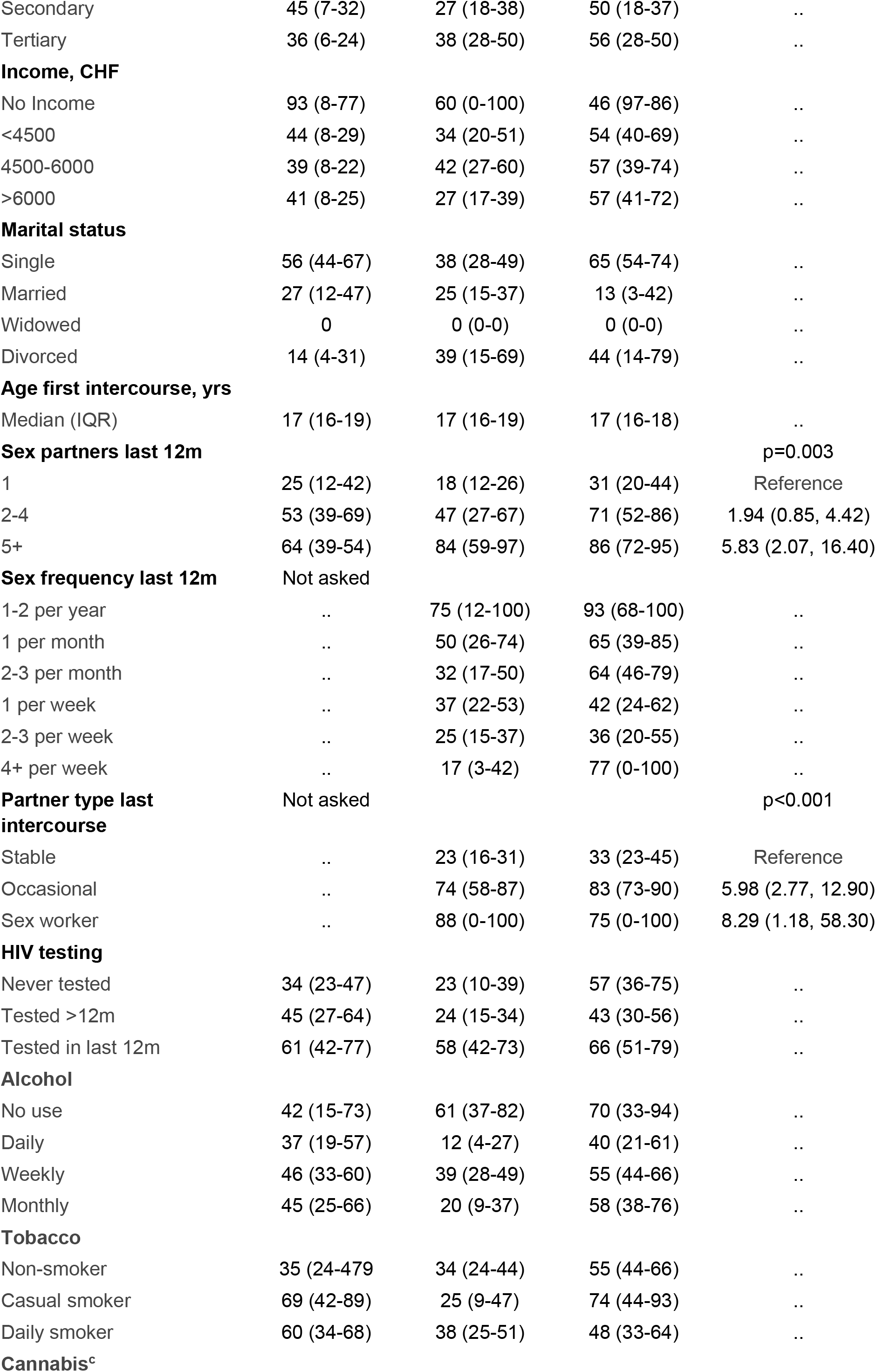
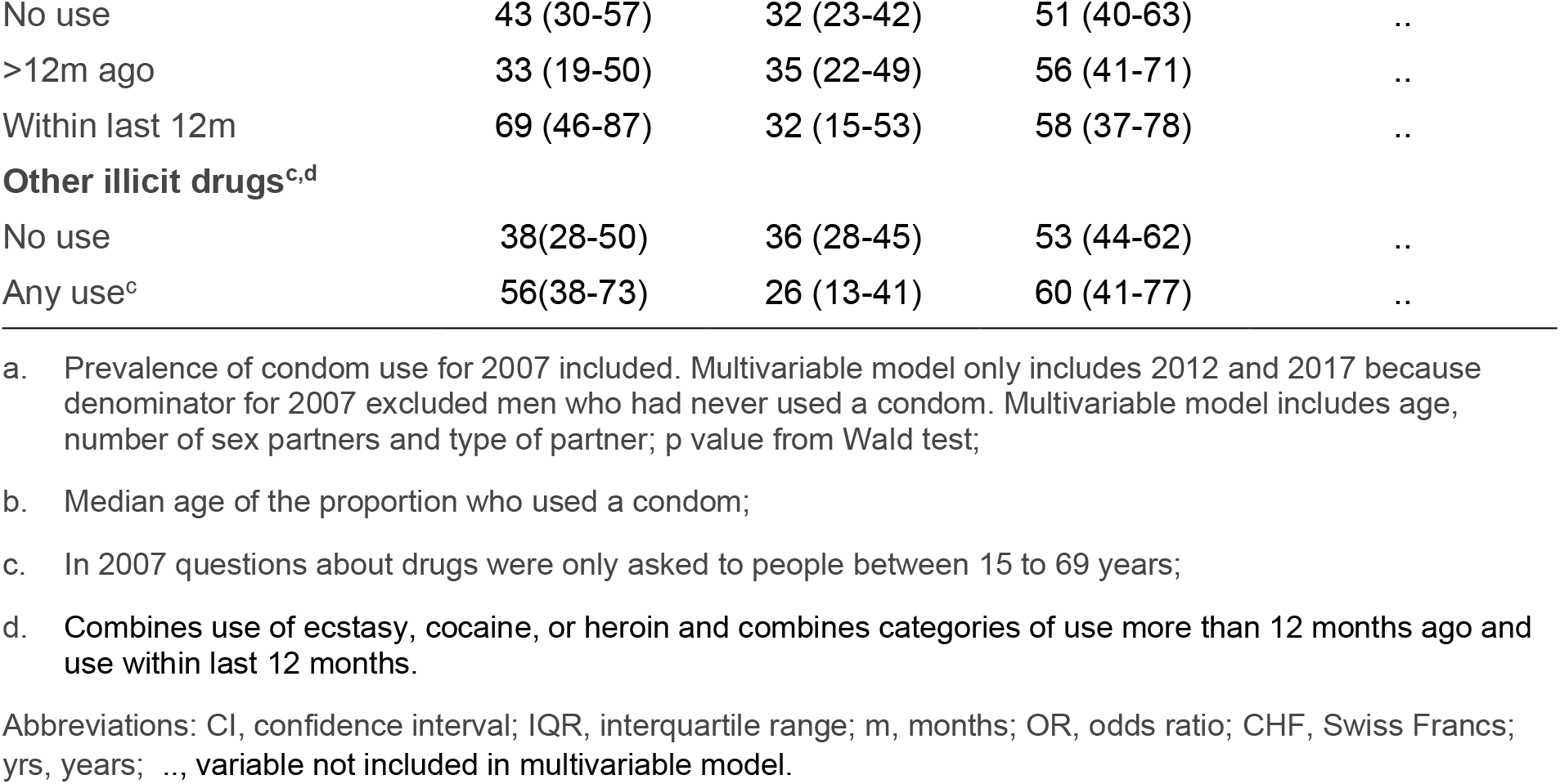
Prevalence of condom use at last sexual intercourse in men reporting any male sex partner, 2007, 2012, 2017 and associations with sociodemographic and behavioural characteristics, 2012 and 2017

The multivariable regression model compared survey years 2012 and 2017 (logistic regression model results for 2007, unadjusted ORs 2012 and 2017, Table S7). There was weak statistical evidence that condom use at last intercourse had increased among men reporting any male sexual partner between 2012 and 2017 (aOR 1.80, 95% CI 0.97, 3.34, p=0.06), after controlling for age, number of sexual partners and type of sexual partner. In the multivariable model, condom use remained strongly associated with higher numbers of sexual partners and for sex with occasional and sex worker partners (aORs 5 or higher, compared with reference groups, unadjusted ORs Table S7).

#### Women reporting any same sex partner

In the group of women who reported any female partner, most reported that they had also had male sex partners (Table S2) and patterns of condom use at last sexual intercourse were similar to those for women with only male partners (Tables S6, S8).

### Testing for HIV infection

#### Men and women reporting only opposite-sex partners

Table 4 shows the lifetime prevalence of having had at least one test for HIV in men and women reporting only opposite-sex partners. For both men and women, respondents in age groups 25 to 44 years were most likely to report ever having had a test for HIV. In the youngest age group, the proportion reporting an HIV test increased considerably in men (29% in 2007, 40% in 2017) but less in women (33% in 2007, 35% in 2017). The highest levels of lifetime HIV testing were reported by those with the highest level of education (in 2017, 52% of men, 64% of women) and those with the highest personal income (in 2017, 52% of men, 61% of women). People who were divorced (in 2017, 61% of men, 66% of women) and who reported 5 or more partners in the last 12 months (in 2017, 52% of men, 74% of women) also reported high levels of having had an HIV test. People who reported use of cannabis (in 2017, 58% of men, 70% of women) and of other illicit drugs (in 2017 66% of men, 83% of women) were more likely than non-users to have had an HIV test (Table S9).

**Table 4.** Prevalence of lifetime HIV testing and associations from multivariable logistic regression, by sociodemographic and behavioural characteristics, in men and women reporting only opposite-sex partners 2007, 2012 and 2017

| HIV testing | Men |  |  |  | Women |  |  |  |
| --- | --- | --- | --- | --- | --- | --- | --- | --- |
|  | 2007 | 2012 | 2017 | Multivariable model<sup>a</sup> | 2007 | 2012 | 2017 | Multivariable model<sup>a</sup> |
|  | Prevalence (95% CI) | Prevalence (95% CI) | Prevalence (95% CI) | Adjusted OR (95% CI) | Prevalence (95% CI) | Prevalence (95% CI) | Prevalence (95% CI) | Adjusted OR (95% CI) |
| **Year** |  |  |  | p<0.001 |  |  |  | p<0.001 |
| 2007 | 32 (30-33) | - | - | Reference | 35 (34-36) | - | - | Reference |
| 2012 | - | 42 (40-43) | - | 1.42 (1.27,1.59) | - | 46 (45-48) | - | 1.35 (1.21,1.50) |
| 2017 | - | - | 45 (44-46) | 1.57 (1.42,1.74) | - | - | 51 (50-53) | 1.54 (1.39,1.71) |
| **Age, years** |  |  |  | p<0.001 |  |  |  | p<0.001 |
| Median (IQR)<sup>b</sup> | 39 (31-49) | 42 (32-50) | 43 (31-47) | 0.99 (0.98,0.99) | 33 (28-38) | 34 (30-38) | 35 (32-39) | 0.97 (0.96,0.97) |
| 16-24 | 20 (17-24) | 29 (25-33) | 40 (37-44) | .. | 56 (52-59) | 63 (60-67) | 62 (59-65) | .. |
| 25-34 | 50 (46-54) | 52 (48-56) | 47 (43-50) | .. | 55 (52-58) | 63 (60-66) | 70 (68-73) | .. |
| 35-44 | 42 (39-45) | 55 (52-58) | 57 (54-60) | .. | 32 (29-35) | 49 (46-52) | 58 (55-60) | .. |
| 45-54 | 33 (30-36) | 44 (41-47) | 53 (50-56) | .. | 15 (12-17) | 22 (19-25) | 38 (35-41) | .. |
| 55-64 | 21 (18-24) | 33 (29-35) | 37 (34-40) | .. | 6 (5-8) | 11 (08-15) | 22 (19-25) | .. |
| 65-74 | 12 (9-14) | 21 (18-25) | 24 (21-27) | .. | 33 (28-38) | 34 (30-38) | 35 (32-39) | .. |
| **Highest educational level** |  |  |  | p<0.001 |  |  |  | p<0.001 |
| No school/primary | 21 (17-25) | 29 (25-33) | 32 (29-36) | Reference | 19 (17-23) | 27 (24-31) | 35 (32-39) | Reference |
| Secondary | 29 (27-31) | 41 (38-43) | 41 (39-43) | 1.74 (1.49,2.03) | 35 (33-36) | 44 (42-46) | 47 (45-49) | 2.07 (1.80,2.39) |
| Tertiary | 40 (37-42) | 47 (45-49) | 52 (50-54) | 2.46 (2.08,2.91) | 52 (49-56) | 60 (58-63) | 64 (62-66) | 3.33 (2.84,3.91) |
| **Income, CHF** |  |  |  | p<0.001 |  |  |  | p<0.001 |
| No Income | 20 (13-29) | 30 (22-38) | 45 (39-51) | 0.75 (0.60,0.93) | 31 (28-35) | 43 (39-47) | 43 (39-47) | 0.91 (0.81,1.02) |
| <4500 | 28 (26-31) | 39 (36-42) | 40 (37-42) | Reference | 34 (33-36) | 45 (43-47) | 45 (43-47) | Reference |
| 4500-6000 | 33 (30-36) | 43 (40-46) | 44 (42-47) | 1.26 (1.13,1.41) | 49 (45-53) | 57 (53-62) | 57 (53-62) | 1.41 (1.23,1.60) |
| >6000 | 37 (35-40) | 46 (43-49) | 52 (50-54) | 1.56 (1.39,1.75) | 56 (49-62) | 61 (55-66) | 61 (55-66) | 1.64 (1.39,1.94) |
| **Marital status** |  |  |  | p<0.001 |  |  |  | p<0.001 |
| Single | 38 (35-41) | 46 (44-49) | 49 (46-51) | 1.17 (1.49,2.03) | 45 (42-48) | 51 (48-54) | 58 (55-60) | 0.69 (0.61,0.78) |
| Married | 26 (25-28) | 37 (35-38) | 40 (38-41) | Reference | 31 (29-33) | 41 (39-43) | 45 (44-47) | Reference |
| Widowed | 19 (11-30) | 41 (27-55) | 43 (29-58) | 1.51 (0.96,2.37) | 16 (12-21) | 35 (24-46) | 33 (26-40) | 1.09 (0.78,1.52) |
| Divorced | 51 (46-56) | 56 (50-61) | 61 (57-66) | 2.38 (2.03,2.79) | 47 (43-51) | 64 (59-69) | 66 (62-70) | 2.29 (1.97,2.66) |
| **Age first intercourse, years** |  |  |  | p<0.001 |  |  |  | p<0.001 |
| Median age (IQR)<sup>b</sup> | 18 (17-19) | 18 (17-19) | 17 (16-18) | 0.95 (0.94,0.97) | 18 (17-19) | 17 (16-19) | 17 (16-19) | 0.96 (0.94,0.98) |
| **Sex partners last 12m** |  |  |  | p=0.014 |  |  |  | p=0.8 |
| 1 | 30 (29-32) | 41 (39-42) | 45 (43-46) | Reference | 37 (35-38) | 46 (44-47) | 52 (50-53) | Reference |
| 2-4 | 42 (37-47) | 46 (41-51) | 54 (49-59) | 1.17 (1.00,1.35) | 50 (43-56) | 57 (50-63) | 69 (63-74) | 1.06 (0.89,1.26) |
| 5+ | 48 (39-58) | 54 (45-62) | 57 (48-65) | 1.37 (1.06,1.76) | 40 (21-62) | 64 (38-85) | 74 (51-91) | 0.90 (0.50,1.61) |
| **Condom use last intercourse** |  |  |  | p<0.001 |  |  |  | p<0.001 |
| Used | 35 (33-37) | 42 (40-44) | 45 (43-46) | Reference | 40 (39-42) | 46 (45-48) | 51 (49-52) | Reference |
| Not used | 33 (30-36) | 41 (38-45) | 46 (44-49) | 0.82 (0.73,0.91) | 42 (39-46) | 47 (43-50) | 54 (51-57) | 0.79 (0.71,0.89) |
| **Alcohol** |  |  |  | p=0.01 |  |  |  | p=0.02 |
| No use | 29 (24-35) | 41 (36-47) | 41 (37-45) | Reference | 66 (63-69) | 47 (43-50) | 47 (44-50) | Reference |
| Daily | 30 (27-33) | 40 (37-44) | 38 (35-41) | 0.80 (0.67,0.96) | 76 (72-80) | 40 (34-45) | 39 (34-44) | 0.73 (0.60,0.89) |
| Weekly | 33 (32-35) | 42 (40-44) | 47 (45-49) | 0.80 (0.69,0.94) | 61 (58-63) | 48 (46-50) | 55 (53-57) | 0.89 (0.79,1.00) |
| Monthly | 31 (27-34) | 43 (39-47) | 46 (43-49) | 0.93 (0.78,1.10) | 66 (63-68) | 45 (43-48) | 50 (48-53) | 0.91 (0.80,1.03) |
| **Tobacco** |  |  |  | p=0.5 |  |  |  | p=0.01 |
| Non-smoker | 28 (27-30) | 39 (37-40) | 43 (42-45) | Reference | 33 (32-35) | 44 (42-45) | 49 (47-50) | Reference |
| Casual smoker | 38 (33-42) | 45 (40-50) | 50 (46-54) | 0.99 (0.89,1.14) | 39 (34-45) | 56 (50-61) | 58 (53-62) | 0.94 (0.80,1.11) |
| Daily smoker | 39 (36-42) | 47 (43-50) | 48 (45-50) | 1.06 (0.96,1.18) | 42 (39-46) | 53 (50-56) | 59 (56-62) | 1.17 (1.05,1.30) |
| **Cannabis<sup>c</sup>** |  |  |  | p<0.001 |  |  |  | p<0.001 |
| No use | 27 (25-29) | 35 (33-37) | 38 (37-40) | Reference | 32 (30-33) | 41 (39-43) | 45 (44-46) | Reference |
| Any use | 48 (44-51) | 56 (53-59) | 58 (55-60) | 1.61 (1.46,1.78) | 61 (58-65) | 67 (63-70) | 70 (68-73) | 1.81 (1.61,2.03) |
| **Other illicit drugs<sup>c,d</sup>** |  |  |  | p<0.001 |  |  |  | p<0.001 |
| No use | 36 (35-37) | 40 (39-42) | 43 (42-44) | Reference | 36 (35-37) | 45 (44-47) | 50 (49-51) | Reference |
| Any use | 79 (71-85) | 65 (59-71) | 66 (61-71) | 1.73 (1.44,2.07) | 79 (71-85) | 81 (74-87) | 83 (78-88) | 2.94 (2.18,3.97) |

a. Multivariable model includes all variables in the table and region of residence, p value from Wald test;
b. Median age of the proportion tested for HIV;
c. In 2007 questions about drugs were only asked to people between 15 to 69 years;
d. Combines use of ecstasy, cocaine, or heroin.

Multivariable analysis showed that lifetime HIV testing increased across all survey years; aOR between 2017 with 2007 was 1.57 (95% CI 1.42, 1.74) for men and 1.54 (1.39, 1.71) for women (Table 4). After adjustment for survey year and all other variables examined, strong associations with ever testing for HIV remained for those with tertiary education vs. no, or primary school only (aOR men 2.46, 95% CI 2.08, 2.91, women 3.33, 95% CI 2.84, 3.91), being divorced vs. married (aOR men 2.38, 95% CI 2.03, 2.79, women 2.29, 95% CI 1.97, 2.66) (unadjusted ORs, Table S10). The strength of associations with drug use was attenuated for use of illicit drugs vs. no use for women (aOR 2.94, 95% CI 2.18, 3.97) and men (aOR 1.7 CI 1.4, 2.1) (Table S11).

#### Men reporting any same sex partner

Most men with any male sex partner reported ever having had an HIV test, with 83% (95% CI 77, 89%) in 2017 (Table 1, Table S12). Among those reporting ever having had an HIV test the highest proportions were aged 25-44 years (Table 5). Men in the oldest age group (65 to 74 years) were least likely to have been tested for HIV. In all survey years, men reporting 5 or more partners in the last year had high levels of HIV testing. For other variables, patterns of lifetime HIV testing were inconsistent. In all survey years, those reporting any use of illicit drugs, were more likely than non-users to have tested for HIV. Across survey years, there was weak evidence of an increase in the lifetime prevalence of HIV testing (2017 vs. 2007, aOR 1.85, 95% CI 0.96, 3.55). The proportions reporting an HIV test in the last 12 months increased from 22% (95% CI 15%, 30%) in 2007 to 30% (14%, 38%) in 2012 and 39% (30%, 47%) in 2017 (Table S12). The factors associated with having had an HIV test in the last year were similar to those associated with ever having had an HIV test (Table S13).

**Table 5.** Prevalence of, and associations with, lifetime HIV testing, by sociodemographic and behavioural characteristics in men reporting any male sex partner, 2007, 2012, 2017

| **HIV testing** | **2007** | **2012** | **2017** | **Multivariable model<sup>a</sup>** |
| --- | --- | --- | --- | --- |
|  | **Prevalence**<br>**(95% CI)** | **Prevalence**<br>**(95% CI)** | **Prevalence**<br>**(95% CI)** | **Adjusted OR**<br>**(95% CI)<sup>a</sup>** |
| **Year** |  |  |  | p=0.2 |
| 2007 | 72 (64-80) | - | - | Reference |
| 2012 | - | 80 (74-60) | - | 1.49 (0.82, 2.69) |
| 2017 | - | - | 83 (77-89) | 1.85 (0.96, 3.55) |
| **Age, years** |  |  |  | p=0.3 |
| Median (IQR)<sup>b</sup> | 38 (30-48) | 42 (35-49) | 39 (32-52) | 0.99 (0.97, 1.01) |
| 16-24 | 67 (00-100) | 43 (14-77) | 62 (38-82) | .. |
| 25-34 | 82 (64-92) | 92 (76-97) | 92 (69-98) | .. |
| 35-44 | 77 (61-88) | 88 (72-95) | 81 (62-92) | .. |
| 45-54 | 64 (38-83) | 89 (78-95) | 96 (85-99) | .. |
| 55-64 | 61 (36-81) | 72 (53-86) | 81 (61-92) | .. |
| 65-74 | 66 (30-90) | 34 (14-63) | 42 (08-87) | .. |
| **Highest educational level** |  |  |  |  |
| No school/primary | 73 (00-100) | 44 (19-73) | 83 (52-96) | .. |
| Secondary | 70 (57-80) | 79 (67-87) | 75 (63-85) | .. |
| Tertiary | 75 (61-85) | 87 (79-92) | 90 (80-95) | .. |
| **Income, CHF** |  |  |  |  |
| No Income | 54 (0-100) | 76 (0-100) | 66 (26-92) | .. |
| <4500 | 57 (42-72) | 74 (58-85) | 75 (60-86) | .. |
| 4500-6000 | 40 (25-58) | 89 (79-95) | 92 (81-97) | .. |
| >6000 | 41 (26-58) | 87 (78-93) | 93 (82-97) | .. |
| **Marital Status** |  |  |  |  |
| Single | 80 (70-88) | 86 (78-92) | 81 (72-88) | .. |
| Married | 47 (30-65) | 75 (64-83) | 82 (53-95) | .. |
| Widowed | - | 100 | - | .. |
| Divorced | 87 (65-96) | 68 (33-90) | 88 (57-98) | .. |
| **Age first intercourse, years** |  |  |  |  |
| Median (IQR) | 17 (15-19) | 17 (16-18) | 17 (16-18) | .. |
| **Sex partners last 12m** |  |  |  | p=0.2 |
| 1 | 68 (53-79) | 77 (68-84) | 82 (70-89) | Reference |
| 2-4 | 73 (57-84) | 83 (66-92) | 91 (80-96) | 1.40 (0.80, 2.45) |
| 5+ | 98 (91-100) | 89 (56-98) | 81 (59-93) | 2.18 (0.86, 5.53) |
| **Condom use last intercourse** |  |  |  |  |
| Used | 69 (56-79) | 77 (68-84) | 84 (74-91) | .. |
| Not used | 83 (71-90) | 87 (75-93) | 83 (72-90) | .. |
| **Alcohol** |  |  |  |  |
| No use | 95 (72-99) | 63 (36-84) | 67 (29-91) | .. |
| Daily | 60 (40-77) | 70 (44-87) | 72 (49-87) | .. |
| Weekly | 76 (64-86) | 84 (77-90) | 84 (74-91) | .. |
| Monthly | 70 (48-85) | 86 (71-94) | 92 (78-97) | .. |
| **Tobacco** |  |  |  |  |
| Non-smoker | 70 (58-80) | 80 (70-87) | 79 (68-87) | .. |
| Casual smoker | 62 (36-82) | 69 (43-87) | 79 (49-94) | .. |
| Daily smoker | 81 (64-91) | 87 (77-93) | 92 (83-97) | .. |
| **Cannabis<sup>c</sup>** |  |  |  |  |
| No use | 59 (45-72) | 70 (59-79) | 84 (74-90) | .. |
| Any use | 82 (71-90) | 91 (84-95) | 83 (70-91) | .. |
| **Other illicit drugs<sup>c, d</sup>** |  |  |  |  |
| No use | 66 (55-75) | 75 (67-82) | 83 (75-88) | .. |
| Any use<sup>c</sup> | 90 (72-97) | 93 (85-97) | 87 (60-96) | .. |

a. Multivariable model includes year, age and number of sex partners; p value from Wald test;
b. Median age of the proportion tested for HIV;
c. In 2007 questions about drugs were only asked to people between 15 to 69 years;
d. Combines use of ecstasy, cocaine, or heroin
Abbreviations: CI, confidence interval; IQR, interquartile range; m, months; OR, odds ratio; CHF, Swiss Francs; .., variable not included in multivariable model.

#### Women reporting any same sex partner

The proportions of women with any female sex partner reporting ever having had an HIV test appeared somewhat higher (61% in 2007, 70% in 2012 and 69% in 2017) than amongst those reporting only male sex partners (35% in 2007, 46 in 2012 and 51% in 2017). There were few consistent patterns according to sociodemographic and behavioural variables (Tables S12, S14). In the multivariable model, there was no strong evidence of an increase across survey years but women reporting higher numbers of sex partners in the last 12 months were most likely to have had an HIV test (5 or more vs. 1 partner, aOR 4.65, 95% CI 1.02, 21.1) (Table S14).

## Discussion

About 1 in 5 men and 1 in 4 women who reported only partners of the opposite sex reported using a condom at last sexual intercourse, with no change from 2012 to 2017. Sex with an occasional rather than a stable partner at last intercourse was the factor most strongly associated with condom use in multivariable analysis. About 1 in 3 to 1 in 2 men who reported any same sex partner said they used a condom at last intercourse with some evidence of an increase from 2012 to 2017 (aOR 1.80, 95% CI 0.97, 3.34). In multivariable analysis, both younger age and higher numbers of sex partners in the last 12 months were strongly associated with condom use. The lifetime prevalence of HIV testing increased from about 1 in 3 to about 1 in 2 from 2007 to 2017 in men and women reporting only partners of the opposite sex. Higher level of education and being divorced were the factors most strongly associated with HIV testing in multivariable analysis. In men with any same sex partner, more than 4 in 5 had ever had a HIV test by 2017. Higher numbers of sex partners remained associated with HIV testing in multivariable analysis. Among women who reported any same sex partner, lifetime prevalence of HIV testing was higher than for women with only opposite-sex partners.

### Strengths and limitations

A strength of this study is that random sampling allows monitoring of condom use and HIV testing amongst a representative population sample that includes men and women who have same sex partners. Surveys among sexual minority groups are usually conducted using convenience sampling methods, which can result in selection biases that tend to overestimate the prevalence of behaviours associated with HIV and STIs.(18) The small numbers of respondents with same sex partners do, however, result in imprecise estimates. Another strength is that we analysed three consecutive surveys although we could only examine changes over time when eligibility criteria for answering the question were similar. We also report data disaggregated by sex and note a small but consistent excess of men who report condom use, even when restricting analyses to people who only report partners of the opposite sex. Since it is the man who wears the condom, this difference might indicate that some women do not consider that they themselves used a condom. A potential limitation of the Swiss Health Survey is that questions were asked during a telephone interview. Direct questioning can result in under-reporting of sensitive behaviours and computer-assisted or written questionnaires are often preferred.(7, 8, 12). If numbers of sexual partners were under-reported and condom use over-reported, odds ratios for the association between condom use and higher sexual partners in the Swiss Health Survey might be over-estimated.

### Interpretation of findings in context of other studies

Direct comparisons of condom use and HIV testing in different national surveys are challenging because definitions of the outcome, the study population and timing of surveys differ in published reports. Overall, condom use at last sexual intercourse in Swiss Health Survey respondents (24% in men and 18% in women reporting only opposite-sex partners aged 16 to 74 years in 2012) appears comparable to the USA General Social Survey (20% of all adults aged 18 years and older from 1996 to 2008) and did not increase over time in either country.(9) In the German Health and Sexuality Survey, the use of a computer-assisted self-interview might have contributed to higher reported use of condoms at last intercourse in single adults aged 18 to 75 years (60% of men, 55% of women in 2018) than in the Swiss Health Survey (48% of men, 38% of women aged 16 to 74 years in 2017).(19) Despite using telephone interviews rather than computer-assisted surveys, the reported prevalence of lifetime HIV testing was higher among adults aged 16 to 74 years in Switzerland (42% of men and 46% of women reporting only opposite-sex partners, 80% of men with any same sex partner in 2012) than in Britain (18% of all men, 23% of all women(10) and 60% of men with any male partner(18) in Natsal 3, conducted from 2010 to 2012).

An advantage of this analysis of data from the Swiss Health Survey was the inclusion of both condom use and HIV testing in the same study. Factors associated with lifetime HIV testing in the Swiss Health Survey in multivariable analysis were more often socioeconomic, whereas sexual behaviours remained associated with condom use. For respondents with only opposite-sex partners, neither condom use at last intercourse nor HIV testing in the last year increased over time. Patterns of condom use in multivariable analyses in the Swiss Health Survey appear broadly consistent with other countries, being more common in the youngest adults, those who are single, with higher numbers of sex partners and with a non-stable partner.(7, 8, 12, 19) Use of illicit drugs was also associated with HIV testing, which might reflect the higher risk of acquiring HIV through shared injection equipment.

Our study provides useful findings about the sexual health of a nationally representative sample of people with same-sex partners. Among men, there is evidence of increases over time in both condom use at last intercourse and lifetime HIV testing, although confidence intervals around the estimates are wide. The proportion who had an HIV test in the last 12 months also increased over time, indicating that the increase was not simply an increase in the cumulative total.(20) Among women who reported any female sex partner, we found similar levels of reported condom use at last intercourse as among women reporting only opposite-sex partners, showing that women with same-sex partners can also be at risk of HIV and STIs. This finding results from decision to group together the small total number of respondents with same-sex partners (Table S2). Most women who reported any same-sex partners also reported having had one or more partners of the opposite sex, so the reported condom use is assumed to have been with a man. Amongst studies that report on condom use by women with same-sex partners, most stratify by self-defined sexual orientation.(11, 12) In the Swiss Health Survey, we could not compare survey responses over time according to sexual orientation because this question was first asked in 2017.

## Conclusions

This study provides data about preventive behaviours, which can be used during the development of the national sexual health programme in Switzerland(21) and comparing outcomes over time and with other countries. Changes over time in levels of condom use and uptake of HIV testing can indicate a need for intensified or targeted prevention information or health promotion, particularly among key population groups at high risk of acquiring or transmitting HIV or other STIs. The number of new diagnoses of HIV infection in Switzerland and other countries in which PrEP use is established is declining,(22) which is consistent with an increase in recent HIV testing in the last 12 months among men seeking PrEP.(5) Further research is warranted to investigate reasons for HIV testing among men with same-sex partners. Reported bacterial STIs in Switzerland are increasing, mostly among men who have sex with men, but also among men and women recorded as heterosexual. An observed increase in condom use among men with same-sex partners is encouraging because the increasing use of PrEP has been associated with an increase in both condomless anal intercourse and increases in reported diagnoses of STIs.(22, 23) Fieldwork for a new round of the Swiss Health Survey took place in 2022. Monitoring of levels of condom use and HIV testing need to continue and changes according to sexual orientation will be possible in future. Although condom use is reported more frequently by people reporting higher numbers of sexual partners, stronger promotion of condoms for people with opposite-sex partners might be needed, since condom use at last intercourse has not changed among since 2007.

## Supporting information

STROBE checklist

## Data Availability

All data produced for this study are available from the Swiss Federal Statistical Office

https://www.bfs.admin.ch/bfs/de/home/statistiken/gesundheit/erhebungen/sgb.html

## List of abbreviations

STI: sexually transmitted infection
CI: Confidence Interval
aOR: adjusted odds ratios
IQR: interquartile range
CHF: Swiss Francs
yrs: years

## Declarations

### Ethics approval and consent to participate

This study did not require approval from a research ethics committee. The data were collected in accordance with the Swiss Federal Statistics Act and were anonymised before sharing. all methods were carried out in compliance with relevant guidelines.

### Availability of data and materials

The data supporting the conclusions of this study are available at the Swiss Federal Statistical Office (https://www.bfs.admin.ch/bfs/de/home/statistiken/gesundheit/erhebungen/sgb.html)

### Author’s contributions

Conceptualisation: NL

Methodology and data curation: DB, NL, GS

Analysis: DB, NL, GS

Writing original draft: DB, NL

Writing review and editing: DB, NL, GS

## Acknowledgments

Marcel Zwahlen advised on the rationale for, and use of, the sample weights provided by the Swiss Federal Office of Statistics for the analysis of the Swiss Health Survey data. Hannah Tough provided valuable comments on earlier drafts of the manuscript.

