## Supplementary material for "Condom use and HIV testing among adults in Switzerland: repeated national cross-sectional surveys 2007, 2012, and 2017"

*Table S1. Swiss Health Survey filtering questions to determine eligible respondents to questions about condom use at last intercourse and HIV testing, according to survey year.*

| Year | Criteria to determine respondents to question "Did you use a male condom the last time you had sex?" |
|---|---|
| 2007 | a. Age between 16 and 74 years<br>b. Reported ever having had sexual intercourse<br>c. Reported ever having used a male condom |
| 2012 | a. Age between 16 and 74 years.<br>b. Reported ever having had sexual intercourse<br>c. Reported ever having had sex during the last 12 months |
| 2017 | a. Age between 16 and 74 years<br>b. Reported ever having had sexual intercourse |
| | **Criteria to determine proportion of people who have had been tested for HIV in their lifetime, question "How many times have you been tested for HIV?" "When was the last time?"** |
| 2007 | a. Age between 16 and 74 years.<br>b. Blood donors: apart from donating blood the number of HIV tests done or<br>c. Non-blood donors: the number of HIV tests done.<br>d. Year and month of first and last test. |
| 2012 | a. Participants older than 16 years<br>b. Blood donors: apart from donating blood the number of HIV tests done or<br>c. Non-blood donors: the number of HIV tests done.<br>d. Year and month the last test. |
| 2017 | a. Participants between 16 and 74 years.<br>b. Number of HIV tests done in life<br>c. Tests done within 12 months or more than 12 months ago. |

*Table S2. Type of partner in men and women reporting any same-sex partner 2007, 2012, 2017*

| Sexual partners | 2007 | 2012 | 2017 |
|---|---|---|---|
| | n (%) | n (%) | n (%) |
| **Men** | | | |
| **Total** | 204 (32) | 246(39) | 183 (29) |
| Mainly with women, but at least one man | 94 (36) | 117 (46) | 47 (18) |
| As many women as men | 20 (37) | 23 (43) | 11 (20) |
| Mainly men but, at least one woman | 43 (44) | 43 (44) | 12 (12) |
| Only with men | 47 (21) | 63 (28) | 113 (51) |
| **Women** | | | |
| **Total** | 227 (32) | 300 (43) | 172 (25) |
| Mainly with men, but at least one woman | 173 (34) | 250(48) | 92 (18) |
| As many men as woman | 22 (51) | 17 (40) | 4 (9) |
| Mainly women but, at least one man | 25 (45) | 23 (41) | 8 (14) |
| Only with women | 7 (8) | 10 (12) | 68 (80) |

*Table S3. Condom use at last sexual intercourse, men and women with only opposite-sex partners. Denominators, unweighted and weighted, for each survey and variable*

| | Men | Men | Men | Women | Women | Women |
|---|---|---|---|---|---|---|
| | 2007 | 2012 | 2017 | 2007 | 2012 | 2017 |
| **Totals, N unweighted, weighted[a]** | 5415, 1972621 | 7382, 2427588 | 8049, 2724669 | 5851,1830947 | 7025, 2132003 | 8428, 2557958 |
| **Age, years** | | | | | | |
| 16-24 | 546, 292091 | 966, 295120 | 903, 301043 | 517, 246538 | 851, 255515 | 876, 258899 |
| 25-34 | 952, 382973 | 1,064, 454261 | 1111, 528079 | 1140, 384853 | 1130, 419164 | 1291, 505920 |
| 35-44 | 1448, 478905 | 1,470, 488511 | 1448, 530176 | 1610, 464059 | 1571, 471335 | 1625, 509388 |
| 45-54 | 1038, 379974 | 1,705, 550479 | 1864, 573081 | 1107, 361793 | 1784, 504578 | 1960, 558324 |
| 55-64 | 874, 275281 | 1,267, 390680 | 1520, 465839 | 947, 253357 | 1086, 307860 | 1604, 434936 |
| 65-74 | 557, 163398 | 910, 248536 | 1203, 326452 | 530, 120346 | 603, 173550 | 1072, 290492 |
| **Region** | | | | | | |
| Lake Geneva | 1019, 369139 | 1317, 440506 | 1424, 508007 | 1089, 343679 | 1290, 407290 | 1550, 479185 |
| Midland | 1408, 444030 | 1435, 542392 | 1579, 587768 | 1555, 424820 | 1389, 472081 | 1739, 593088 |
| Northwest | 575, 260440 | 1026, 325278 | 849, 352834 | 631, 253925 | 979, 291493 | 904, 351691 |
| Zurich | 753, 365687 | 740, 415267 | 880, 488010 | 756, 316728 | 663, 344353 | 921, 443955 |
| East | 593, 282339 | 1117, 369950 | 1528, 390425 | 589, 233062 | 994, 300103 | 1487, 341979 |
| Central | 694, 179458 | 1218, 229857 | 1227, 275223 | 806, 187883 | 1187, 217857 | 1252, 241036 |
| Ticino | 373, 71528 | 529, 104338 | 562, 122401 | 425, 70849 | 523, 98827 | 575, 107024 |
| **Highest educational level** | | | | | | |
| No school/primary | 481, 215272 | 842, 274694 | 951, 292971 | 701, 230403 | 894, 275746 | 1035, 288781 |
| Secondary | 2940, 1057960 | 3599, 1181527 | 3783, 1264045 | 3931, 1223891 | 4315, 1283197 | 4772, 1400556 |
| Tertiary | 1993, 699184 | 2924, 966760 | 3301, 1163362 | 1218, 376452 | 1797, 567319 | 2600, 860448 |
| Missing | 1, 206 | 17, 4608 | 14, 4291 | 1, 201 | 19, 5740 | 21, 8173 |
| **Income, SFr** | | | | | | |
| No income | 139, 60317 | 25, 177565 | 346, 112507 | 679, 222076 | 918, 279067 | 978, 271987 |
| <4500 | 1772, 679214 | 2344, 790486 | 2689, 918762 | 3350, 1091013 | 4031, 1221547 | 4953, 1466363 |
| 4500-6000 | 1357, 465421 | 1718, 570588 | 1897, 654101 | 746, 210379 | 730, 222686 | 1049, 356014 |
| >6000 | 1617, 576518 | 2242, 725455 | 2524, 850147 | 349, 95689 | 461, 132703 | 735, 251417 |
| Missing | 530, 191151 | 827, 263494 | 593, 189153 | 727, 211789 | 885, 276000 | 713, 212177 |
| **Marital status** | | | | | | |
| Single | 1762, 697087 | 2281, 849328 | 2407, 995481 | 1678, 554665 | 1798, 585228 | 2255, 795789 |
| Married | 2915, 1084242 | 4391, 328466 | 4916, 1447463 | 3011, 1017360 | 4400, 1262541 | 5105, 1414870 |
| Widowed | 97, 21835 | 8428, 715 | 71, 24841 | 314, 62188 | 132, 40937 | 206, 59874 |
| Divorced | 639, 168110 | 575, 219908 | 655, 256884 | 846, 196433 | 693, 241865 | 862, 287426 |
| Missing | 2, 1347 | 3, 1171 | 0,0 | 2, 301 | 2, 1431 | 0,0 |
| **Sex partners last 12m** | | | | | | |
| 1 | 4191, 1558594 | 6382, 2095192 | 6498, 2158653 | 4649, 1548175 | 6583, 1995736 | 6960, 2094370 |
| 2-4 | 665, 244120 | 731, 243845 | 666, 255717 | 410, 124385 | 397, 121682 | 374, 127432 |
| 5+ | 197, 74241 | 228, 77697 | 199, 79313 | 34, 10711 | 29, 8387 | 30, 11663 |
| Missing | 362, 95668 | 41, 10854 | 686, 230987 | 758, 147675 | 16, 6198 | 1064, 324494 |
| **Sex frequency last 12m** | Not asked | | | Not asked | | |
| 1-2 per year | .. | 271, 80731 | 351, 118565 | .. | 325, 93051 | 376, 111423 |
| 1 per month | .. | 828, 261277 | 1028, 329464 | .. | 864, 249163 | 1124, 338486 |
| 2-3 per month | .. | 1472, 473009 | 1701, 546318 | .. | 1283, 375995 | 1718, 511495 |
| 1 per week | .. | 1991, 650719 | 1944, 665376 | .. | 1975, 607513 | 2090, 629519 |
| 2-3 per week | .. | 2267, 765183 | 1962, 693239 | .. | 2071, 652754 | 1680, 531014 |
| 4+ per week | .. | 398, 149006 | 320, 118262 | .. | 257, 79908 | 210, 70530 |

| | | | | | | |
|---|---|---|---|---|---|---|
| Missing | .. | 155, 47662 | 743, 253443 | .. | 250, 73619 | 1219, 365490 |
| **Partner type last intercourse** | Not asked | | | Not asked | | |
| Stable | .. | 6665, 2190882 | 7173, 2393191 | .. | 6714, 2040553 | 7982, 2409173 |
| Occasional | .. | 677, 226436 | 803, 302565 | .. | 306, 90071 | 433, 145050 |
| Sex worker | .. | 31, 7646 | 67, 26208 | .. | 0, 0 | 1, 200 |
| Missing | .. | 9, 2585 | 6, 2705 | .. | 5, 1380 | 12, 3535 |
| **HIV testing** | | | | | | |
| Never tested | 3432, 1660169 | 4345, 1383746 | 4580, 1482996 | 4679, 1499320 | 3831, 1128432 | 4344, 1228391 |
| >12m ago | 439, 202837 | 2337, 801418 | 2857, 1017787 | 778, 222246 | 2443, 767295 | 3432, 1096425 |
| Within last 12m | 203, 93129 | 487, 171075 | 524, 197190 | 953, 98305 | 568, 182702 | 537, 199462 |
| Missing | 43, 16488 | 213, 71350 | 88, 26695 | 41, 11078 | 183, 53574 | 115, 33680 |
| **Alcohol** | | | | | | |
| No use | 403, 145469 | 582, 218019 | 818, 285192 | 936, 284968 | 1132, 342310 | 1533, 470954 |
| Daily | 1041, 332620 | 1261, 375407 | 1175, 352970 | 470, 130237 | 549, 152674 | 521, 143233 |
| Weekly | 3010, 1135926 | 4171, 1362093 | 4626, 1583047 | 2432, 791798 | 3137, 958014 | 3963, 1228355 |
| Monthly | 957, 356529 | 1367, 471796 | 1428, 502750 | 2012, 623797 | 2206, 678812 | 2410, 715234 |
| Missing | 4, 2078 | 1, 273 | 2,711 | 1, 146 | 1, 193 | 1, 182 |
| **Tobacco** | | | | | | |
| Non-smoker | 3528, 1279407 | 4891, 1575101 | 5488, 1807651 | 4235, 1338398 | 5175, 1557199 | 6315, 1889971 |
| Casual smoker | 550, 203001 | 726, 244321 | 776, 287648 | 415, 134856 | 486, 147355 | 605, 196721 |
| Daily smoker | 1337, 490213 | 1764, 606873 | 1785, 629371 | 1199, 357248 | 1364, 427449 | 1507, 471044 |
| Missing | - | 7, 1293 | 0, 0 | 2, 444 | 0, 0 | 1, 222 |
| **Cannabis**[b] | | | | | | |
| No use | 3525, 1284285 | 5302, 1693509 | 5549, 1, 770092 | 4457, 1405317 | 5828, 1740879 | 6547, 1912407 |
| >12m ago | 1302, 485363 | 1525, 548295 | 1836, 683783 | 1035, 334662 | 959, 314830 | 1532, 523148 |
| Within last 12m | 311, 123641 | 534, 178092 | 633, 258879 | 114, 36826 | 221, 71061 | 333, 118541 |
| Missing | 277, 7933 | 21, 7693 | 31,11915 | 245, 54142 | 17, 5233 | 13, 3863 |
| **Other illicit drugs**[b, c] | | | | | | |
| No use | 4837, 1788061 | 6958, 2275033 | 7469, 2479592 | 5441, 1725723 | 6798, 2058121 | 8123, 2444996 |
| >12m ago | 308, 105962 | 392, 140247 | 522, 217782 | 176, 54453 | 217, 70348 | 289, 106705 |
| Within last 12m | 13, 4404 | 22, 8031 | 45, 21281 | 2, 754 | 7, 2877 | 12, 5399 |
| Missing | 257, 74195 | 10, 4277 | 13, 6013 | 232, 50016 | 3, 656 | 4, 858 |

a. Overall denominators for condom use and HIV testing differ because eligibility criteria for answering each question differed;

b. In 2007 questions about drugs were only asked to people between 15 to 69 years;

c. Combines use of ecstasy, cocaine, or heroin

Abbreviations: m, months; SFr, Swiss Francs; yrs, years; .., question not asked.

*Table S4. Condom use, men and women reporting only opposite-sex partners, 2007. Prevalence and associations with sociodemographic factors, sexual behaviours and substance use*

| Condom Use | Men 2007 | | | Women 2007 | | |
|---|---|---|---|---|---|---|
| | **Prevalence (95% CI)** | **Unadjusted OR (95% CI)** | **Adjusted OR (95% CI)[a]** | **Prevalence (95% CI)** | **Unadjusted OR (95% CI)** | **Adjusted OR (95% CI) [a]** |
| **Total** | 26 (24-27) | | | 20 (18-21) | | |
| **Age, years** | | p<0.001 | p<0.001 | | p<0.001 | p<0.001 |
| Median (IQR)[b] | 18 (16-19) | 0.93 (0.93, 0.94) | 0.96 (0.95, 0.97) | 18 (16-20) | 0.95 (0.94, 0.95) | 0.95 (0.94,0.96) |
| 16-24 | 65 (60-70) | - | - | 43 (38-49) | - | - |
| 25-34 | 30 (27-34) | - | - | 25 (22-28) | - | - |
| 35-44 | 21 (19-24) | - | - | 18 (16-21) | - | - |
| 45-54 | 14 (11-18) | - | - | 12 (10-15) | - | - |
| 55-64 | 9 (7-12) | - | - | 7 (5-10) | - | - |
| 65-74 | 9 (7-13) | - | - | 6 (5-10) | - | - |
| **Region** | | p=0.9 | | | p=0.015 | p=0.10 |
| Lake Geneva | - | reference | reference | - | reference | reference |
| Midland | 25 (22-28) | 0.94 (0.74,1.20) | 0.98 (0.73, 1.33) | 19 (16-21) | 1.00 (0.78, 1.28) | 1.31 (0.95, 1.79) |
| Northwest | 25 (21-30) | 0.95 (0.71,1.26) | 1.02 (0.72, 1.45) | 16 (13-19) | 0.82 (0.61, 1.11) | 0.98 (0.67, 1.44) |
| Zurich | 25 (22-29) | 0.94 (0.73,1.21) | 1.11 (0.80, 1.55) | 23 (20-27) | 1.31 (1.01, 1.71) | 1.53 (1.08, 2.17) |
| East | 27 (23-32) | 1.05 (0.78,1.41) | 0.89 (0.62, 1.29) | 21 (17-26) | 1.18 (0.86, 1.61) | 1.54 (1.04, 2.27) |
| Central | 25 (20-29) | 0.92 (0.68, 1.23) | 0.92 (0.64, 1.33) | 23 (19-27) | 1.27 (0.96, 1.68) | 1.31 (0.91, 1.89) |
| Ticino | 27 (22-33) | 1.06 (0.77, 1.45) | 1.07 (0.69, 1.65) | 16 (12-20) | 0.82 (0.58, 1.16) | 1.05 (0.66, 1.67) |
| **Highest education level** | | p<0.001 | p=0.2 | | p<0.001 | p=0.030 |
| No school/primary | 47 (41-53) | reference | reference | 26 (22-31) | reference | reference |
| Secondary | 26 (23-28) | 0.39 (0.30, 0.50) | 0.71 (0.50, 1.02) | 18 (16-19) | 0.61 (0.47, 0.78) | 0.65 (0.47, 0.91) |
| Tertiary | 19 (17-21) | 0.27 (0.20, 0.35) | 0.71 (0.47, 1.06) | 21 (19-24) | 0.75 (0.56, 1.00) | 0.76 (0.52, 1.12) |
| **Income, SFr** | | p<0.001 | p=0.10 | | p=0.6 | p=0.9 |
| No Income | 48(45-51) | 2.19 (1.42, 3.38) | | 38 (35-40) | 0.87 (0.66, 1.15) | 1.08 (0.79, 1.47) |
| <4500 | 13 (11-14) | reference | reference | 10 (9-11) | reference | reference |
| 4500-6000 | 6 (3-11) | 0.46 (0.37, 0.56) | 0.76 (0.59, 0.98) | 15 (10-22) | 1.06 (0.84, 1.34) | 0.98 (0.73, 1.33) |
| >6000 | 20 (16-24) | 0.31 (0.25, 0.39) | 0.75 (0.56, 1.01) | 19 (16-23) | 0.90 (0.64, 1.27) | 1.12 (0.73, 1.73) |
| **Marital status** | | p<0.001 | p<0.001 | | p<0.001 | p<0.001 |
| Single | 55 (45-65) | 6.44 (5.34, 7.76) | 2.02 (1.53, 2.66) | 18 (15-22) | 5.40 (4.46, 6.55) | 2.10 (1.58, 2.80) |
| Married | 36 (33-39) | reference | reference | 20 (19-22) | reference | reference |
| Widowed | 20 (18-23) | 0.44 (0.21, 0.91) | 0.16 (0.04, 0.63) | 22 (18-25) | 1.63 (1.03, 2.60) | 2.07 (0.98, 4.38) |
| Divorced | 15 (13-17) | 1.72 (1.29, 2.31) | 1.27 (0.87, 1.86) | 19 (14-24) | 2.11 (1.61, 2.77) | 1.61 (1.10, 2.36) |
| **Age first intercourse, yrs** | | p= 0.004 | p<0.001 | | p=0.3 | p<0.001 |
| Median (IQR)[b] | 18 (16-19) | 0.96 (0.93, 0.99) | 1.1 (1.06, 1.14) | 18 (16.0, 20.0) | 0.98 (0.95, 1.01) | 1.11 (1.07, 1.15) |
| <16 | 31 (27-35) | - | - | 23 (20-27) | - | - |
| 16-18 | 25 (22-27) | - | - | 20 (18-22) | - | - |
| 19-25 | 23 (21-25) | - | - | 17 (15-19) | - | - |
| 26-30 | 19 (12-27) | - | - | 20 (13-30) | - | - |
| >30 | 27 (8-53) | - | - | 30 (10-60) | - | - |
| **Sex partners last 12m** | | p<0.001 | p<0.001 | | p<0.001 | p<0.001 |
| 1 | 18 (16-19) | reference | reference | 15 (14-17) | reference | reference |
| 2-4 | 54 (49-59) | 5.39 (4.28, 6.79) | 2.89 (2.17, 3.85) | 50 (44-56) | 5.45 (4.17, 7.11) | 2.79 (2.05, 3.80) |
| 5+ | 65 (57-74) | 8.68 (5.87, 12.8) | 3.7 (2.31, 5.94) | 40 (20-63) | 3.73 (1.54, 9.03) | 3.02 (1.22, 7.50) |
| **HIV testing** | | p=0.8 | p=0.6 | | p=0.016 | p=0.7 |
| Never | 26 (24-27) | reference | reference | 19 (18-21) | reference | reference |
| >12m ago | 25 (21-30) | 0.99 (0.77, 1.26) | 1.09 (0.79, 1.48) | 20 (17-24) | 1.06 (0.84, 1.33) | 1.11 (0.83, 1.48) |
| Within last 12m | 28 (21-35) | 1.11 (0.79, 1.57) | 0.84 (0.55, 1.29) | 27 (22-33) | 1.56 (1.15, 2.11) | 1.08 (0.74, 1.58) |
| **Alcohol** | | p<0.001 | p=0.14 | | p<0.001 | p=0.6 |
| No use | 33 (26-40) | reference | reference | 18 (15-22) | reference | reference |
| Daily | 14 (11-16) | 0.31 (0.21, 0.46) | 0.58 (0.37, 0.92) | 10 (7-14) | 0.50 (0.32,0.77) | 0.98 (0.55, 1.76) |
| Weekly | 27 (25-30) | 0.77 (0.56, 1.08) | 0.74 (0.50, 1.10) | 20 (18-22) | 1.13 (0.88, 1.46) | 1.02 (0.73, 1.42) |
| Monthly | 28 (24-32) | 0.79 (0.55, 1.13) | 0.75 (0.49, 1.15) | 22 (20-24) | 1.26 (0.97, 1.62) | 1.18 (0.84, 1.64) |
| **Tobacco** | | p=0.044 | p=0.4 | | p=0.008 | p=0.065 |

| | | | | | | |
|---|---|---|---|---|---|---|
| Non-smoker | 24 (22-26) | reference | reference | 19 (17-20) | reference | reference |
| Casual smoker | 27 (22-32) | 1.17 (0.89, 1.53) | 0.94 (0.68, 1.30) | 26 (22-32) | 1.56 (1.17,2.06) | 0.93 (0.64, 1.37) |
| Daily smoker | 29 (26-32) | 1.26 (1.04, 1.51) | 0.83 (0.65, 1.07) | 20 (17-23) | 1.09 (0.89, 1.33) | 0.71 (0.53, 0.95) |
| **Cannabis**[c] | | p<0.001 | p=0.8 | | p<0.001 | p=0.5 |
| No use | 23 (21-25) | reference | reference | 18 (17-20) | reference | reference |
| >12m ago | 31 (28-34) | 1.51 (1.25, 1.81) | 0.96 (0.75, 1.24) | 24 (21-28) | 1.42 (1.16, 1.75) | 0.98 (0.74, 1.31) |
| Within last 12m | 45 (38-52) | 2.83 (2.09, 3.82) | 1.09 (0.72, 1.67) | 39 (29-50) | 2.86 (1.82, 4.49) | 1.37 (0.79, 2.36) |
| **Other illicit drugs**[c,d] | | p=0.059 | p=0.8 | | p<0.001 | p=0.017 |
| No use | 26 (25-27) | reference | reference | 20 (18-21) | reference | reference |
| >12m ago | 33 (27-40) | 1.43 (1.04, 1.96) | 0.88 (0.58, 1.34) | 35 (27-44) | 2.17 (1.47, 3.21) | 2.18 (1.26, 3.61) |
| Within last 12m | 38 (8-77) | 1.75 (0.50, 6.14) | 1.03 (0.14, 7.56) | 49 (1-100) | 3.88 (0.24, 62.2) | 1.90 (0.21, 17.2) |

a. Multivariable model includes all variables in the table, p value from Wald test;

b. Median age of the proportion who used a condom;

c. In 2007 questions about drugs were only asked to people between 15 to 69 years;

d. Combines use of ecstasy, cocaine, or heroin.

Abbreviations: CI, confidence interval; m, months; OR, odds ratio; SFr, Swiss Francs; yrs, years.

*Table S5. Condom use at last sexual intercourse, men and women with only opposite-sex partners, 2012 and 2017. Univariable logistic regression*

| | **Men 2012** | **Men 2017** | **Women 2012** | **Women 2017** |
|---|---|---|---|---|
| **Condom use** | **Unadjusted OR (95% CI)** [a] | **Unadjusted OR (95% CI)**[a] | **Unadjusted OR (95% CI)** [a] | **Unadjusted OR (95% CI)** [a] |
| **Age, yrs** | p<0.001 | p<0.001 | p<0.001 | p<0.001 |
| | 0.92 (0.92, 0.93) | 0.94 (0.93, 0.94) | 0.93 (0.92, 0.93) | 0.94 (0.94, 0.95) |
| **Region** | p=0.4 | p=0.8 | p=0.13 | p=0.009 |
| Lake Geneva | reference | reference | reference | reference |
| Midland | 0.88 (0.71, 1.10) | 0.89 (0.74, 1.08) | 1.33 (1.04, 1.71) | 0.75 (0.62, 0.91) |
| Northwest | 0.94 (0.74, 1.18) | 0.94 (0.75, 1.17) | 1.25 (0.95, 1.64) | 0.97 (0.78, 1.21) |
| Zurich | 0.96 (0.75, 1.23) | 0.98 (0.79, 1.21) | 1.36 (1.03, 1.79) | 0.97 (0.78, 1.20) |
| East | 0.93 (0.71, 1.21) | 0.9 (0.74, 1.10) | 1.29 (0.97, 1.72) | 1 (0.82, 1.23) |
| Central | 1.11 (0.88, 1.39) | 1.03 (0.83, 1.28) | 1.44 (1.12, 1.84) | 0.96 (0.76, 1.21) |
| Ticino | 0.79 (0.60, 1.05) | 0.96 (0.75, 1.23) | 1.33 (0.97, 1.82) | 0.68 (0.52, 0.89) |
| **Highest education level** | p<0.001 | p<0.001 | p=0.025 | p<0.001 |
| No school/primary | reference | reference | reference | reference |
| Secondary | 0.59 (0.48, 0.72) | 0.64 (0.54, 0.77) | 0.79 (0.62, 1.01) | 0.76 (0.63, 0.92) |
| Tertiary | 0.4 (0.33, 0.50) | 0.64 (0.54, 0.77) | 0.98 (0.74, 1.28) | 0.99 (0.81, 1.21) |
| **Income, SFr** | p<0.001 | p<0.001 | p<0.001 | p=0.3 |
| No income | 2.62 (1.85, 3.69) | 1.91 (1.46, 2.49) | 1.19 (0.95, 1.47) | 1.23 (1.02, 1.49) |
| <4500 | reference | reference | reference | reference |
| 4500-6000 | 0.44 (0.36, 0.54) | 0.56 (0.47, 0.66) | 0.91 (0.69, 1.20) | 1.1 (0.91, 1.34) |
| >6000 | 0.33 (0.28, 0.40) | 0.45 (0.38, 0.52) | 0.96 (0.69, 1.34) | 1.01 (0.80, 1.28) |
| **Marital status** | p<0.001 | p<0.001 | p<0.001 | p<0.001 |
| Single | 6.73 (5.72, 7.91) | 6.37 (5.54, 7.32) | 4.92 (4.15, 5.84) | 4.25 (3.69, 4.90) |
| Married | reference | reference | reference | reference |
| Widowed | 0.97 (0.47, 2.03) | 0.98 (0.38, 2.50) | 0.96 (0.48, 1.95) | 0.81 (0.47, 1.39) |
| Divorced | 1.5 (1.10, 2.05) | 1.6 (1.23, 2.07) | 1.22 (0.87, 1.70) | 1.68 (1.35, 2.10) |
| **Age first intercourse, yrs** | p<0.001 | p=0.3 | p<0.001 | p=0.9 |
| Per year increase | 0.9 (0.88, 0.93) | 0.99 (0.98, 1.01) | 0.93 (0.90, 0.96) | 1 (0.98, 1.02) |
| **Sex partners last 12m** | p<0.001 | p<0.001 | p<0.001 | p<0.001 |
| 1 | reference | reference | reference | reference |
| 2-4 | 8.78 (7.03, 11.0) | 6.55 (5.33, 8.05) | 6.92 (5.32, 9.00) | 5.71 (4.46, 7.33) |
| 5+ | 12.3 (7.97, 19.1) | 11.6 (7.84, 17.2) | 8.71 (3.38, 22.4) | 5.81 (2.45, 13.8) |
| **Sex frequency last 12 m** | p<0.001 | p<0.001 | p<0.001 | p<0.001 |
| 1-2 per year | 2.53 (1.76, 3.63) | 1.78 (1.32, 2.39) | 2.24 (1.53, 3.28) | 1.96 (1.45, 2.66) |
| 1 per month | reference | reference | reference | reference |
| 2-3 per month | 0.81 (0.62, 1.04) | 0.85 (0.69, 1.04) | 0.94 (0.72, 1.23) | 0.83 (0.67, 1.04) |
| 1 per week | 0.47 (0.36, 0.61) | 0.62 (0.51, 0.77) | 0.66 (0.51, 0.86) | 0.74 (0.59, 0.91) |
| 2-3 per week | 0.54 (0.42, 0.70) | 0.45 (0.36, 0.55) | 0.62 (0.48, 0.80) | 0.7 (0.55, 0.87) |
| 4+ per week | 0.66 (0.45, 0.96) | 0.56 (0.39, 0.81) | 0.51 (0.27, 0.98) | 0.68 (0.44, 1.04) |
| **Partner type last intercourse** | p<0.001 | p<0.001 | p<0.001 | p<0.001 |
| Stable | reference | reference | reference | reference |
| Occasional | 20.8 (15.8, 27.5) | 12.2 (9.98, 14.9) | 14 (10.1, 19.4) | 9.46 (7.37, 12.1) |
| Sex worker | 26.1 (9.52, 71.4) | 26.3 (11.1, 62.8) | - | - |
| **HIV testing** | p<0.001 | p<0.001 | p<0.001 | p<0.001 |
| Never tested | reference | reference | reference | reference |
| >12m ago | 0.84 (0.71, 0.99) | 0.95 (0.84, 1.09) | 0.91 (0.77, 1.08) | 1.03 (0.90, 1.17) |
| Within last 12m | 1.9 (1.45, 2.50) | 1.77 (1.41, 2.22) | 1.58 (1.23, 2.04) | 1.83 (1.45, 2.31) |
| **Alcohol** | p<0.001 | p=0.001 | p<0.001 | p<0.001 |
| No use | reference | reference | reference | reference |
| Daily | 0.37 (0.26, 0.52) | 0.28 (0.21,0.38) | 0.42 (0.27, 0.65) | 0.35 (0.24,0.52) |
| Weekly | 0.86 (0.65, 1.14) | 0.83 (0.68,1.01) | 1.33 (1.04, 1.69) | 0.87 (0.74, 1.04) |
| Monthly | 0.88 (0.64, 1.20) | 1.06 (0.84, 1.32) | 1.67 (1.30, 2.15) | 1.02 (0.84, 1.22) |
| **Tobacco** | p=0.11 | p=0.11 | p<0.001 | p<0.001 |
| Non-smoker | reference | reference | reference | reference |
| Casual smoker | 1.44 (1.12, 1.86) | 1.24 (1.01, 1.51) | 1.68 (1.29, 2.19) | 1.79 (1.45, 2.22) |
| Daily smoker | 1.27 (1.07, 1.49) | 1.02 (0.88, 1.19) | 1.21 (0.99, 1.48) | 1.11 (0.94, 1.31) |
| **Cannabis** | p<0.001 | p<0.001 | p<0.001 | p<0.001 |
| No use | reference | reference | reference | reference |
| >12m ago | 1.17 (0.97, 1.41) | 1.15 (0.99, 1.34) | 1.42 (1.15, 1.75) | 1.28 (1.09, 1.49) |
| Within last 12m | 2.97 (2.35, 3.74) | 1.99 (1.61, 2.44) | 3.17 (2.17, 4.63) | 2.41 (1.83, 3.16) |
| **Other illicit drugs**[b] | p<0.001 | p<0.001 | p=0.016 | p=0.075 |
| No use | reference | reference | reference | reference |

| | | | | |
|---|---|---|---|---|
| >12m ago | 1.48 (1.11, 1.97) | 1.24 (0.98, 1.57) | 1.62 (1.09, 2.41) | 1.15 (0.82, 1.60) |
| Within last 12m | 5.12 (1.66, 15.7) | 1.63 (0.84, 3.17) | 3.89 (0.75, 20.3) | 4.41 (1.17, 16.6) |

a. p value from Wald test;

b. Combines use of ecstasy, cocaine, or heroin.

Abbreviations: CI, confidence interval; m, months; NA, not applicable; OR, odds ratio; SFr, Swiss Francs; yrs, years.

*Table S6. Condom use at last sexual intercourse, men and women with any same-sex partners. Denominators, unweighted and weighted, and number of missing values for each survey and variable*

| Condom use | Men | Men | Men | Women | Women | Women |
|---|---|---|---|---|---|---|
| | **2007** | **2012** | **2017** | **2007** | **2012** | **2017** |
| **Totals, N unweighted, weighted[a]** | 191, 66118 | 245, 84238 | 183, 76438 | 205, 63179 | 300, 96257 | 172, 69018 |
| **Age, years** | | | | | | |
| 16-24 | 7, 3497 | 20, 5643 | 25, 8197 | 18, 8593 | 55, 15295 | 35, 14084 |
| 25-34 | 46, 18408 | 34, 14478 | 32, 17291 | 49, 17849 | 80, 31574 | 55, 25382 |
| 35-44 | 63, 21425 | 68, 27822 | 40, 18722 | 63, 15055 | 66, 20741 | 30, 12090 |
| 45-54 | 35, 10938 | 62, 19514 | 38, 16023 | 53, 16743 | 65, 18851 | 36, 12713 |
| 55-64 | 27, 8762 | 40, 11544 | 37, 13623 | 17, 3800 | 24, 6718 | 10, 3117 |
| 65-74 | 13, 3089 | 21, 5235 | 11, 2581 | 5, 1139 | 10, 3077 | 6, 1631 |
| **Region** | | | | | | |
| Lake Geneva | 30,11415 | 45, 15147 | 46, 19171 | 54, 14541 | 64, 19810 | 43, 16336 |
| Midland | 50,16811 | 36, 15951 | 18, 8757 | 43, 11187 | 53, 18694 | 38, 18342 |
| Northwest | 18,6636 | 44, 11119 | 20, 11332 | 29, 10918 | 59, 18221 | 16, 6521 |
| Zurich | 48,19165 | 51, 24919 | 38, 19435 | 43, 17835 | 41, 21542 | 26, 14769 |
| East | 20,6109 | 28, 9991 | 27, 10121 | 15, 4411 | 31, 8353 | 20, 6475 |
| Central | 14,3722 | 34, 5939 | 26, 5903 | 17, 3850 | 37, 6536 | 20, 4442 |
| Ticino | 11,2261 | 7, 1170 | 8, 1718 | 4, 437 | 15, 3100 | 9, 2133 |
| **Highest educational level** | | | | | | |
| No school/ primary | 8, 3424 | 19, 4915 | 16, 5477 | 10, 3162 | 28, 8517 | 17, 5036 |
| Secondary | 101, 31318 | 111, 39221 | 83, 32537 | 121, 39253 | 160, 54460 | 83, 33636 |
| Tertiary | 82, 31377 | 115, 40102 | 84, 38423 | 74, 20764 | 112, 33279 | 72, 30346 |
| Missing | 0, 0 | 0, 0 | 0, 0 | 0, 0 | 0, 0 | 0, 0 |
| **Income, SFr** | | | | | | |
| No Income | 5, 2205 | 9, 2488 | 12, 3984 | 14, 5114 | 20, 6896 | 8, 2581 |
| <4500 | 72, 26242 | 70, 22070 | 64, 26740 | 111, 34980 | 184, 59501 | 106, 44156 |
| 4500-6000 | 48, 16155 | 49, 16453 | 45, 19462 | 41, 12314 | 39, 11864 | 31, 10373 |
| >6000 | 50, 17086 | 85, 32340 | 50, 20141 | 23, 5677 | 29, 9141 | 22, 9965 |
| Missing | 16, 4432 | 32, 10885 | 12, 6110 | 16, 5,094 | 28, 8854 | 5, 1943 |
| **Marital status** | | | | | | |
| Single | 126, 42994 | 121, 46054 | 122, 52867 | 111, 32720 | 149, 49696 | 111, 47453 |
| Married | 38, 13661 | 101, 28417 | 18, 5042 | 42, 16386 | 105, 29705 | 22, 7883 |
| Widowed | 0, 0 | 1, 178 | 0, 0 | 3, 634 | 5, 1679 | 2, 428 |
| Divorced | 26, 8350 | 22, 9589 | 14, 5571 | 49, 13439 | 40, 14608 | 14, 5518 |
| Missing | 1, 1113 | 0, 0 | 29, 12956 | 0, 0 | 1, 569 | 23, 7735 |
| **Age first intercourse, yrs** | | | | | | |
| <16 | 59, 23916 | 94, 34200 | 70, 29812 | 89, 29114 | 162, 50085 | 76, 30797 |
| 16-18 | 63, 94439 | 80, 28715 | 64, 26995 | 71, 23506 | 87, 29698 | 53, 22093 |
| 19-25 | 60, 19581 | 64, 19584 | 44, 17456 | 42, 10261 | 44, 14199 | 40, 14872 |
| 26-30 | 3, 853 | 3, 746 | 3, 1175 | 1, 69 | 1, 183 | 2, 1003 |
| >30 | 1, 183 | 0, 0 | 0, 0 | 2, 230 | 2, 830 | 0, 0 |
| Missing | 5, 1613 | 2, 992 | 1, 998 | 0, 0 | 2, 1261 | 1, 254 |
| **Sex partners last 12m** | | | | | | |
| 1 | 72,27206 | 163, 53952 | 89, 36364 | 125, 42000 | 241, 79105 | 106, 44848 |
| 2-4 | 67,22612 | 48, 18743 | 41, 19755 | 35, 10162 | 45, 13384 | 41, 16743 |
| 5+ | 29,10790 | 29, 10335 | 38, 15068 | 14, 3568 | 14, 3767 | 7, 3005 |
| Missing | 23, 5511 | 4, 1208 | 15, 5250 | 31, 7448 | 0, 0 | 18, 2506 |
| **Sex frequency last 12m** | **Not asked** | | | **Not asked** | | |
| 1-2 per year | .. | 11, 3497 | 13, 3918 | .. | 22, 6591 | 8, 3402 |
| 1 per month | .. | 27, 7807 | 26, 10429 | .. | 32, 10029 | 29, 11156 |
| 2-3 per month | .. | 51, 16847 | 43, 19311 | .. | 54, 19159 | 32, 12295 |

| | | | | | | |
|---|---|---|---|---|---|---|
| 1 per week | .. | 58, 16937 | 41, 18896 | .. | 69, 21662 | 38, 15715 |
| 2-3 per week | .. | 76, 33157 | 39, 16550 | .. | 83, 27004 | 31, 15140 |
| 4+ per week | .. | 17, 4665 | 7, 2827 | .. | 33, 10111 | 16, 6888 |
| Missing | .. | 5, 1327 | 14, 4504 | .. | 7, 1701 | 18, 4422 |
| **Partner type, last intercourse** | **Not asked** | | | **Not asked** | | |
| Stable | .. | 188, 67115 | 102, 43535 | .. | 255, 84285 | 137, 54869 |
| Occasional | .. | 53, 16300 | 76, 31278 | .. | 45, 11972 | 33, 13418 |
| Sex worker | .. | 4, 822 | 5, 1625 | .. | 0, 0 | 0, 0 |
| Missing | .. | 0, 0 | 0, 0 | .. | 0, 0 | 2, 731 |
| **HIV testing** | | | | | | |
| Never tested | 94, 32314 | 61, 16837 | 37, 12357 | 129, 39530 | 81, 27912 | 56,21757 |
| >12m ago | 49, 17309 | 111, 40139 | 80, 33188 | 45, 13132 | 158, 48215 | 80, 33248 |
| Within last 12m | 44, 15623 | 66, 24807 | 61, 28525 | 28, 9144 | 52, 16840 | 36, 14012 |
| Missing | 4, 871 | 7, 2454 | 5, 2367 | 3, 1373 | 9, 3289 | 0, 0 |
| **Alcohol** | | | | | | |
| No use | 18, 4430 | 28, 8021 | 13, 4645 | 18, 6690 | 33, 10778 | 24, 10064 |
| Daily | 37, 13811 | 37, 12283 | 30, 10228 | 24, 6480 | 24, 7291 | 11, 4582 |
| Weekly | 99, 33535 | 137, 48188 | 101, 46129 | 109, 32512 | 161, 49932 | 93, 36322 |
| Monthly | 37, 14343 | 43, 15746 | 39, 15436 | 53, 17245 | 82, 28256 | 44, 18050 |
| Missing | 0, 0 | 0, 0 | 0, 0 | 1, 252 | 0, 0 | 0, 0 |
| **Tobacco** | | | | | | |
| Non-smoker | 104, 38210 | 141, 48931 | 106, 45271 | 97, 28610 | 143, 49064 | 84, 34367 |
| Casual smoker | 23, 6326 | 31, 11905 | 18, 5952 | 20, 6154 | 51, 16581 | 19, 7859 |
| Daily smoker | 64, 21582 | 73, 23402 | 59, 25215 | 88, 28415 | 106, 30611 | 69, 26792 |
| Missing | 0, 0 | 0, 0 | 0, 0 | 0, 0 | 0, 0 | 0, 0 |
| **Cannabis**[b] | | | | | | |
| No use | 91, 26776 | 129, 42310 | 94, 38959 | 79, 23164 | 103, 33352 | 74, 28862 |
| >12m ago | 63, 27025 | 74, 25507 | 56, 25133 | 91, 28213 | 133, 44220 | 65, 26828 |
| Within last 12m | 29, 9899 | 40, 15510 | 31, 11225 | 32, 10254 | 64, 18685 | 31, 12594 |
| Missing | 8, 2418 | 2, 910 | 2, 1121 | 3, 1547 | 0 | 2, 734 |
| **Other illicit drugs**[b, c] | | | | | | |
| No use | 142, 47328 | 185, 60328 | 148, 62088 | 154, 47431 | 234, 75008 | 136, 53648 |
| Any use | 42, 16651 | 57, 23152 | 35, 14350 | 51, 15748 | 66, 21248 | 36, 15370 |
| Missing | 7, 2140 | 3, 757 | 0, 0 | 0, 0 | 0, 0 | 0, 0 |

a. Overall denominators for condom use and HIV testing differ because eligibility criteria for answering each question differed;

b. In 2007 questions about drugs were only asked to people between 15 to 69 years;

c. Combines use of ecstasy, cocaine, or heroin and combines categories of use more than 12 months ago and use within last 12 months.

Abbreviations: m, months; SFr, Swiss Francs; yrs, years; .., question not asked

*Table S7. Condom use, men reporting any same-sex partner. Associations with sociodemographic factors, sexual behaviours and substance use from univariable logistic regression, 2007, 2012, 2017 and from multivariable logistic regression, 2007.*

| **Condom Use** | **2007** | **2007** | **2012** | **2017** |
|---|---|---|---|---|
| | **Unadjusted OR (95% CI)** | **Adjusted OR (95% CI)** [a] | **Unadjusted OR (95% CI)** | **Unadjusted OR (95% CI)** |
| **Age, years** | p<0.001 | p=0.057 | p<0.001 | p<0.001 |
| **Per year increase** | 0.97 (0.94-1.00) | 0.97 (0.93, 1.00) | 0.95 (0.92, 0.97) | 0.95 (0.93, 0.98) |
| **Region** | p=0.9 | | p=0.4 | P=0.9 |
| Lake Geneva | reference | | reference | reference |
| Midland | 1.46 (0.44, 4.85) | | 0.62 (0.20, 1.98) | 0.99 (0.29, 3.32) |
| Northwest | 0.78 (0.18, 3.34) | | 0.86 (0.30, 2.49) | 1.12 (0.35, 3.55) |
| Zurich | 1.19 (0.39, 3.59) | | 1.69 (0.68, 4.21) | 0.86 (0.33, 2.21) |
| East | 1.09 (0.26, 4.59) | | 1.5 (0.44, 5.04) | 0.85 (0.27, 2.64) |
| Central | 3.04 (0.51, 18.0) | | 0.65 (0.21, 1.95) | 1.47 (0.49, 4.42) |
| Ticino | 0.95 (0.14, 6.36) | | 2.06 (0.32, 13.4) | 1.09 (0.21, 5.71) |
| **Highest education level** | p=0.013 | | p=0.2 | p=0.5 |
| No school/primary | reference | | reference | reference |
| Secondary | 0.11 (0.02, 0.66) | | 0.41 (0.10, 1.71) | 0.51 (0.14, 1.82) |
| Tertiary | 0.07 (0.01, 0.44) | | 0.7 (0.17, 2.84) | 0.64 (0.18, 2.29) |
| **Income, SFr** | p=0.11 | | p=0.2 | P=>0.9 |
| No Income | 16.5 (1.43, 191) | | 2.88 (0.53, 15.5) | 0.72 (0.18, 2.93) |
| <4500 | reference | | reference | reference |
| 4500-6000 | 0.81 (0.31, 2.08) | | 1.41 (0.54, 3.72) | 1.1 (0.44, 2.76) |
| >6000 | 0.87 (0.35, 2.16) | | 0.71 (0.29, 1.73) | 1.1 (0.48, 2.55) |
| **Marital Status** | p.001 | | p=<0.001 | p=0.001 |
| Single | 3.33 (1.25-8.87) | | 1.88 (0.89, 3.98) | 12.4 (3.00, 51.2) |
| Married | reference | | reference | reference |
| Widowed | NA | | - | - |
| Divorced | 0.43 (0.12, 1.60) | | 2.12 (0.59, 7.69) | 5.3 (0.82, 34.1) |
| **Age first sexual intercourse, years** | p=0.8 | | p=0.2 | p=0.4 |
| **Median (IQR)** | 0.99 (0.89, 1.09) | | 0.93 (0.84, 1.05) | 1.04 (0.94, 1.16) |
| **Sex partners 12m** | p=0.015 | p=0.014 | p<0.001 | p<0.001 |
| 1 | reference | reference | reference | reference |
| 2-4 | 3.45 (1.28, 9.28) | 3.73 (1.40, 9.92) | 3.86 (1.51, 9.87) | 5.55 (2.02, 15.2) |
| 5+ | 5.31 (1.50, 18.7) | 4.70 (1.30, 16.9) | 22.6 (5.56, 91.9) | 13.4 (4.62, 38.6) |
| **Average sex frequency 12 m** | Not asked | | p=0.030 | p=0.006 |
| 1-2 times per year | .. | | 3.10 (0.54, 18.0) | 7.55 (1.08, 52.5) |
| Once per month | .. | | reference | reference |
| 2-3 per month | .. | | 0.47 (0.14, 1.55) | 0.96 (0.29, 3.15) |
| Once per week | .. | | 0.58 (0.19, 1.83) | 0.4 (0.12, 1.38) |
| 2-3t/week | .. | | 0.33 (0.11, 1.00) | 0.31 (0.09, 1.05) |
| 4>/week | .. | | 0.20 (0.04, 0.92) | 1.78 (0.20, 15.8) |
| **Type of partner last sex intercourse** | Not asked | | p<0.001 | p<0.001 |
| Stable partner | .. | | reference | reference |
| Occasional partner | .. | | 9.85 (4.14, 23.5) | 9.81 (4.46, 21.6) |
| Sex-worker | .. | | 25.1 (2.35, 269) | 5.94 (0.61, 57.5) |
| **HIV Testing** | p=0.051 | | p<0.001 | p<0.001 |
| Never tested | reference | | reference | reference |
| Tested >2m | 1.57 (0.63, 3.88) | | 1.08 (0.41, 2.87) | 0.57 (0.22, 1.47) |
| Tested in last 12m | 2.95 (1.22, 7.09) | | 4.76 (1.70, 13.3) | 1.46 (0.55, 3.90) |
| **Alcohol** | p=0.9 | | p=0.001 | p=0.3 |
| No use | reference | | reference | reference |
| Daily | 0.8 (0.21, 3.03) | | 0.09 (0.02, 0.34) | 0.29 (0.07, 1.28) |
| Weekly | 1.15 (0.33, 3.97) | | 0.4 (0.15, 1.10) | 0.53 (0.14, 2.03) |
| Monthly | 1.11 (0.28, 4.43) | | 0.16 (0.05, 0.56) | 0.6 (0.14, 2.61) |
| **Tobacco** | p= 0.031 | | p=0.6 | p=0.2 |
| Non-smoker | reference | | reference | reference |
| Casual smoker | 4.09 (1.32, 12.6) | | 0.66 (0.22, 1.95) | 2.26 (0.71, 7.19) |
| Daily smoker | 1.89 (0.82, 4.40) | | 1.2 (0.59, 2.41) | 0.76 (0.36, 1.63) |
| **Cannabis**[b] | p= 0.031 | | p=0.9 | p=0.8 |
| No use | reference | | reference | reference |
| >12m ago | 2.9 (1.01, 8.34) | | 1.14 (0.55, 2.38) | 1.22 (0.56-2.68) |
| Within last 12m | 0.64 (0.27, 1.53) | | 1.01 (0.38, 2.64) | 1.32 (0.51-3.39) |
| **Other illicit drugs**[b, c] | p= 0.089 | | p=0.2 | p=0.5 |
| No use | reference | | reference | reference |
| Any use[b] | 2.09 (0.89, 4.91) | | 0.61 (0.27, 1.37) | 1.31 (0.58,3.10) |

a. Multivariable model for 2007, includes age and number of sex partners; p value from Wald test;

b. In 2007 questions about drugs were only asked to people between 15 to 69 years;

c. Combines use of ecstasy, cocaine, or heroin and combines categories of use more than 12 months ago and use within last 12 months.

Abbreviations: CI, confidence interval; m, months; OR, odds ratio; SFr, Swiss Francs; yrs, years.

*Table S8. Condom use, women reporting any same-sex partner. Associations with sociodemographic factors, sexual behaviours and substance use from univariable logistic regression, 2007, 2012, 2017 and from multivariable logistic regression 2007 and multivariable logistic regression 2012, 2017.*

| **Condom Use** | **2007** | | | **2012** | | **2017** | | |
|---|---|---|---|---|---|---|---|---|
| | **Prevalence (95 CI%)** | **Unadjusted OR (95% CI)** | **Adjusted OR (95% CI)** [a] | **Prevalence (95 CI%)** | **Unadjusted OR (95% CI)** | **Prevalence (95 CI%)** | **Unadjusted OR (95% CI** | **Adjusted OR (95% CI)** [a] |
| | 23 (17-31) | | | | | | | |
| **Year** | | | | | | | | 0.084 |
| 2012 | .. | | | 27 (21-33) | | .. | | reference |
| 2017 | .. | | | .. | | 25 (18-33) | | 0.59 (0.32, 1.08) |
| **Age, years** | | p=0.058 | p=0.2 | | p=0.009 | | p<0.001 | p=0.011 |
| Median (IQR)[b] | 31 (24-43) | 0.96(0.93-1.00) | 0.97 (0.92, 1.02) | 30 (22-41) | 0.96 (0.93, 0.99 | 26 (23-31) | 0.92 (0.88, 0.96) | 0.96 (0.94, 0.98) |
| 16-24 | 51 (22-79) | .. | .. | 49 (33-66) | .. | 39 (21-60) | .. | .. |
| 25-34 | 29 (17-45) | .. | .. | 28 (17-42) | .. | 35 (22-51) | .. | .. |
| 35-44 | 13 (7-23) | .. | .. | 22 (12-35) | .. | 18 (7-40) | .. | .. |
| 45-54 | 12 (6-23) | .. | .. | 18 (9-32) | .. | 5 (1-16) | .. | .. |
| 55-64 | 34 (10-71) | .. | .. | 19 (3-51) | .. | 0 (0-0) | .. | .. |
| 65-74 | 20 (0-94) | .. | .. | 6 (0-69) | .. | 11 (0-0) | .. | .. |
| **Highest educational level** | | p=0.7 | | | p=0.6 | | p=0.5 | |
| No school/primary | 16 (0-41) | reference | .. | 37 (16-62) | reference | 38 (10-73) | reference | .. |
| Secondary | 26 (16-36) | 1.87 (0.26, 13.5) | .. | 25 (17-34) | 0.57 (0.21, 1.58) | 27 (17-39) | 0.61 (0.16, 2.30) | .. |
| Tertiary | 21 (11-32) | 1.43 (0.19, 10.6) | .. | 27 (18-38) | 0.64 (0.22, 1.81) | 21 (11-33) | 0.44 (0.11, 1.69) | .. |
| **Income, SFr** | | p=0.11 | | | p=0.025 | | p=0.9 | |
| No Income | 22 (0-47) | 1.00 (0.20, 4.99) | .. | 10 (1-30) | 0.28 (0.07, 1.10) | 35 (3-83) | 1.58 (0.33, 7.64) | .. |
| <4500 | 22 (12-31) | reference | .. | 29 (21-37) | reference | 26 (17-36) | reference | .. |
| 4500-6000 | 24 (9-39) | 1.15 (0.42, 3.12) | .. | 32 (17-52) | 0.57 (0.21, 1.58) | 23 (7-46) | 0.61 (0.16, 2.30) | .. |
| >6000 | 32 (6-57) | 1.66 (0.45, 6.08) | .. | 8 (2-21) | 0.64 (0.22, 1.81) | 21 (6-47) | 0.44 (0.11, 1.69) | .. |
| **Marital Status** | | p<0.001 | | | p<0.001 | | p<0.001 | |
| Single | 34 (23-45) | 4.33 (1.33, 14.1) | .. | 39 (30-49) | 4.36 (2.05, 9.27) | 40 (17-69) | 0.55 (0.17, 1.74) | .. |
| Married | 11 (0-21) | reference | .. | 13 (7-22) | reference | 0 (0-0) | reference | .. |
| Widowed | 0 (0-0) | - | .. | 16 (0-46) | 1.29 (0.12, 13.3) | 11 (2-47) | - | .. |
| Divorced | 15 (5-26) | 1.49 (0.39, 5.73) | .. | 15 (6-35) | 1.39 (0.39, 5.02) | 40 (17-69) | 0.18 (0.03, 1.06) | .. |
| **Age first intercourse, yrs** | | p=0.5 | | | p=0.043 | | p=0.11 | |
| Median (IQR)[b] | 17 (15-18) | 1.05 (0.92, 1.21) | .. | 16 (15-17) | 0.86 (0.74, 0.99) | 16 (14-18) | 0.89 (0.77, 1.03) | .. |
| **Sex partners last 12m** | | p<0.001 | p=0.005 | | p<0.001 | | p<0.001 | p<0.001 |
| 1 | 13 (7-21) | reference | reference | 20 (14-27) | reference | 11 (5-20) | reference | reference |
| 2-4 | 48 (26-70) | 6.31 (2.17, 18.3) | 5.10 (1.61, 16.2) | 61 (43-78) | 6.49 (2.83, 14.9) | 51 (33-70) | 8.25 (2.98, 22.8) | 5.96 (3.12, 11.4) |
| 5+ | 57 (17-90) | 9.06 (2.11, 39.0) | 7.39 (1.47, 37.2) | 56 (17-89) | 5.11 (1.26, 20.7) | 55 (5-97) | 9.35 (1.64, 53.3) | 4.99 (1.58, 15.7) |
| **Sex frequency last 12m** | Not asked | Not asked | | | p=0.005 | | p=0.08 | |
| 1-2 per year | .. | .. | .. | 40 (16-69) | reference | 29 (0-97) | reference | .. |
| 1 per month | .. | .. | .. | 40 (20-62) | 1.03 (0.27, 3.93) | 19 (5-41) | 1.75 (0.20, 15.6) | .. |
| 2-3 per month | .. | .. | .. | 43 (26-61) | 1.13 (0.38, 3.38) | 30 (13-51) | 1.82 (0.44, 7.57) | .. |
| 1 per week | .. | .. | .. | 19 (10-31) | 0.35 (0.12, 1.03) | 31 (15-51) | 1.98 (0.49, 7.99) | .. |
| 2-3 per week | .. | .. | .. | 12 (5-21) | 0.2 (0.07, 0.61) | 17 (5-35) | 0.87 (0.19, 3.97) | .. |
| 4+ per week | .. | .. | .. | 36 (17-57) | 1.03 (0.27, 3.93) | 18 (1-57) | 1.75 (0.20, 15.6) | .. |
| **Partner type last intercourse** | Not asked | Not asked | | | p<0.001 | | p<0.001 | |
| Stable | .. | .. | .. | 20 (15-27) | reference | 15 (9-23) | reference | .. |
| Occasional | .. | .. | .. | 72 (56-85) | 10.2 (4.49, 23.1) | 68 (45-87) | 12.5 (4.12, 38.0) | .. |
| Sex worker | .. | .. | .. | 0 (0-0) | - | 0 (0-0) | - | .. |
| **HIV testing** | | p=0.2 | | | p=0.9 | | p=0.2 | |
| Never tested | 18 (12-27) | reference | .. | 28 (17-41) | reference | 24 (12-39) | reference | .. |
| Tested >12-m | 29 (15-47) | 1.84 (0.75, 4.52) | .. | 29 (20-38) | 1.06 (0.50, 2.23) | 20 (11-32) | 0.82 (0.31, 2.20) | .. |
| Tested in last 12m | 37 (15-65) | 2.64 (0.83, 8.38) | .. | 25 (13-40) | 0.87 (0.34, 2.22) | 39 (21-59) | 2.03 (0.69, 5.95) | .. |
| **Alcohol** | | p=0.6 | | | p=0.6 | | p=0.074 | |
| No use | 24 (2-68) | reference | .. | 24 (10-43) | reference | 9 (2-23) | reference | .. |
| Daily | 14 (3-36) | 0.52 (0.06, 4.50) | .. | 13 (1-44) | 0.46 (0.06, 3.48) | 26 (1-76) | 3.3 (0.46, 23.8) | .. |
| Weekly | 27 (18-38) | 1.19 (0.19, 7.46) | .. | 30 (22-39) | 1.39 (0.53, 3.65) | 32 (22-44) | 4.54 (1.37, 15.0) | .. |
| Monthly | 20 (10-34) | 0.81 (0.12, 5.55) | .. | 25 (14-39) | 1.08 (0.36, 3.27) | 19 (8-35) | 2.25 (0.57, 8.87) | .. |
| **Tobacco** | | p=0.7 | | | p=0.14 | | p=0.022 | |
| Non-smoker | 23 (14-34) | reference | .. | 21 (13-30) | reference | 14 (7-24) | reference | .. |
| Casual smoker | 33 (10-64) | 1.66 (0.49, 5.62) | .. | 37 (22-53) | 2.22 (0.95, 5.21) | 39 (13-70) | 3.79 (1.07, 13.5) | .. |
| Daily smoker | 22 (12-35) | 0.96 (0.41, 2.24) | .. | 32 (21-43) | 1.78 (0.85, 3.72) | 35 (22-49) | 3.21 (1.29, 7.95) | .. |

| | | | | | | | | |
|---|---|---|---|---|---|---|---|---|
| **Cannabis[c]** | | p=0.5 | | | p=0.069 | | p=0.038 | |
| No use | 19 (10-30) | reference | .. | 23 (15-34) | reference | 16 (8-28) | reference | .. |
| >12m ago | 24 (14-37) | 1.36 (0.56, 3.31) | .. | 23 (15-34) | 1 (0.47, 2.13) | 26 (15-39) | 1.82 (0.68, 4.88) | .. |
| Within last 12m | 31 (14-53) | 1.92 (0.65, 5.69) | .. | 41 (27-57) | 2.3 (1.04, 5.06) | 45 (24-67) | 4.27 (1.39, 13.1) | .. |
| **Other illicit drugs[c,d]** | | p<0.001 | | | p=0.2 | | p=0.6 | |
| No use | 34 (16-51) | reference | .. | 25 (18-32) | reference | 42 (16-32) | reference | .. |
| Any used | 20 (13-28) | 1.98 (0.80,4.93) | .. | 35 (20-49) | 1.61 (0.78, 3.34) | 29 (12-46) | 0.48, 3.30) | .. |

a. Multivariable models separate for 2007 and 2012, 2017 because denominator for 2007 excluded women who had never used a condom. Multivariable models include age, number of sex partners and type of partner; p value from Wald test;

b. Median age of the proportion who used a condom;

c. In 2007 questions about drugs were only asked to people between 15 to 69 years;

d. Combines use of ecstasy, cocaine, or heroin and combines categories of use more than 12 months ago and use within last 12 months.

Abbreviations: CI, confidence interval; IQR, interquartile range; m, months; OR, odds ratio; SFr, Swiss Francs; yrs, years; .., variable not included in multivariable model.

*Table S9. Lifetime HIV testing, men and women with only opposite-sex partners. Denominators, unweighted and weighted, for each survey and variable, 2007, 2012, 2017*

| HIV testing | Men | Men | Men | Women | Women | Women |
|---|---|---|---|---|---|---|
| | 2007 | 2012 | 2017 | 2007 | 2012 | 2017 |
| **Totals, N unweighted, weighted[a]** | 6236, 2237900 | 7063, 2313010 | 7975, 2702130 | 7582, 2289504 | 6763, 2049462 | 8330, 2529129 |
| **Age, years** | | | | | | |
| 16-24 | 553, 296239 | 949, 290087 | 901, 300476 | 524, 248000 | 843, 255515 | 874, 258899 |
| 25-34 | 955, 384253 | 1040, 446058 | 1104, 525416 | 1183, 402818 | 1100, 419164 | 1278, 505920 |
| 35-44 | 1506, 505797 | 1402, 465779 | 1436, 525957 | 1753, 521670 | 1511, 471335 | 1604, 509388 |
| 45-54 | 1184, 441374 | 1629, 529873 | 1842, 566707 | 1344, 446311 | 1681, 504578 | 1921, 558324 |
| 55-64 | 1181, 365700 | 1179, 363623 | 1505, 461133 | 1532, 401480 | 1049, 307860 | 1592, 434936 |
| 65-74 | 857, 244536 | 864, 234265 | 1187, 322440 | 1246, 269225 | 579, 173550 | 1061, 290492 |
| **Region** | | | | | | |
| Lake Geneva | 1165, 413941 | 1255, 421571 | 1412, 504218 | 1416, 429093 | 1223, 407290 | 1526, 479185 |
| Midland | 1651, 506532 | 1375, 521007 | 1563, 582248 | 2011, 519597 | 1332, 472081 | 1717, 593088 |
| Northwest | 657, 299390 | 984, 312365 | 842, 350702 | 806, 319638 | 944, 291493 | 889, 351691 |
| Zurich | 812, 398380 | 710, 397487 | 873, 485356 | 935, 386575 | 648, 344353 | 915, 443955 |
| East | 711, 334068 | 1067, 356147 | 1510, 385186 | 789, 304118 | 962, 300103 | 1477, 341979 |
| Central | 806, 202729 | 1175, 222817 | 1222, 273641 | 1032, 233854 | 1157, 217857 | 1240, 241036 |
| Ticino | 1165, 413941 | 1255, 421571 | 1412, 504218 | 593, 96630 | 497, 98827 | 566, 107024 |
| **Highest education level** | | | | | | |
| No school/primary | 613, 255693 | 808, 262368 | 941, 289892 | 1182, 347612 | 857, 275746 | 1019, 288781 |
| Secondary | 3443, 1220851 | 3436, 1132883 | 3756, 1256566 | 5039, 1523082 | 4163, 1283197 | 4726, 1400556 |
| Tertiary | 2179, 761150 | 2803, 930103 | 3264, 1151381 | 1358, 417617 | 1725, 567319 | 2566, 860448 |
| Missing | 1, 206 | 16, 4330 | 14,4291 | 3,1193 | 18, 5740 | 19, 8173 |
| **Income, SFr** | | | | | | |
| No income | 149, 64349 | 244, 75987 | 342, 111526 | 889, 294891 | 888, 279067 | 963, 271987 |
| <4500 | 2134, 789202 | 2253, 759570 | 2660, 910239 | 4367, 1346627 | 3897, 1221547 | 4896, 1466363 |
| 4500-6000 | 1520, 523117 | 1648, 549727 | 1881, 649750 | 862, 241467 | 699, 222686 | 1042, 356014 |
| >6000 | 1769, 630922 | 2154, 697210 | 2503, 842885 | 407, 109029 | 443, 132703 | 732, 251417 |
| Missing | 664, 230310 | 764, 247191 | 589,187729 | 1057,297489 | 836, 276000 | 697, 212177 |
| **Marital status** | | | | | | |
| Single | 1790, 702281 | 2230, 829308 | 2397, 991450 | 1776, 572834 | 1751, 585228 | 2247, 795789 |
| Married | 3600, 1323818 | 4208, 1263895 | 4859, 1430224 | 4065, 1353870 | 4226, 1262541 | 5024, 1414870 |
| Widowed | 149, 31389 | 80, 27635 | 70, 24647 | 684, 122382 | 126, 40937 | 206, 59874 |
| Divorced | 695, 179067 | 543, 208182 | 649, 255809 | 1054, 239916 | 658, 241865 | 853, 287426 |
| Missing | 2, 1347 | 2,664 | 0,0 | 3,502 | 2, 1431 | 0,0 |
| **Sex partners last 12m** | | | | | | |
| 1 | 4883, 1793871 | 6086, 2005936 | 6437, 2140357 | 5811, 1908525 | 6331, 1924121 | 6876,1924121 |
| 2-4 | 666, 242075 | 712, 236900 | 660, 254133 | 404, 123338 | 384, 118102 | 488,118102 |
| 5+ | 200, 76497 | 221, 74953 | 197, 78844 | 34, 10340 | 29, 8387 | 29,8387 |
| Missing | 487, 106440 | 44,11896 | 681, 228796 | 1333, 110513 | 19, 6301 | 1053, 320588 |
| **Condom use last intercourse** | | | | | | |
| Used | 3936, 1244522 | 5424, 1772851 | 6027, 1758067 | 4519, 1057221 | 5567, 1688823 | 6710, 1997371 |
| Not used | 1270, 654750 | 1630, 553994 | 1934, 552103 | 1141, 726368 | 1188, 365366 | 1603, 526907 |
| Missing | 1030, 338630 | 9, 2838 | 15, 4156 | 3234, 505916 | 8, 2721 | 1209, 4850 |
| **Alcohol** | | | | | | |
| No use | 504, 182855 | 557, 208127 | 802, 280558 | 1370, 404354 | 1082, 329781 | 1504, 461747 |
| Daily | 1329, 415464 | 1185, 354083 | 1161, 348790 | 747, 195648 | 526, 146025 | 516, 142503 |
| Weekly | 3301, 1, 235, 3 | 4002, 1310690 | 4589, 1571742 | 2905, 921169 | 3032, 926154 | 3924, 1215081 |
| Monthly | 1097, 401590 | 1318, 456511 | 1421, 500330 | 2558, 767830 | 2122, 654757 | 2385, 500330 |
| Missing | 5, 2653 | 1,273 | 2, 711 | 2, 504 | 1, 93 | 1,182 |

| Tobacco | | | | | | |
|---|---|---|---|---|---|---|
| Non-smoker | 4122, 1465671 | 4675, 1512349 | 5437, 1791473 | 5614, 1703513 | 4994, 1512349 | 6243, 1896416 |
| Casual smoker | 599, 216270 | 694, 233922 | 768, 285872 | 227, 145214 | 463, 233922 | 596, 193855 |
| Daily smoker | 1515, 555960 | 1693, 582120 | 1770, 624782 | 622, 440333 | 1306, 582120 | 1490, 465636 |
| Missing | 0, 0 | 1, 1293 | 0,0 | 2, 444 | 0, 0 | 1,222 |
| **Cannabis**[b] | | | | | | |
| No use | 4203, 1509223 | 4808, 1540934 | 5492, 1753661 | 5797, 1778908 | 5406, 1540934 | 6460, 1887252 |
| Any use | 1612, 614244 | 1985, 702051 | 2453, 936771 | 1184, 384976 | 1133, 702051 | 1857, 638014 |
| Missing | 421, 114433 | 270, 86968 | 30, 11698 | 588, 125620 | 224, 65444 | 13, 3863 |
| **Other illicit drugs**[b,c] | | | | | | |
| No use | 5520, 2020619 | 6663, 2185257 | 7400, 2459983 | 6821, 2113861 | 6549, 2185257 | 8026, 2416588 |
| Any use | 313, 107355 | 393, 141195 | 562, 236133 | 173, 54604 | 212, 141195 | 300, 111683 |
| Missing | 403, 109927 | 7, 3231 | 13, 6013 | 588,121040 | 2, 371 | 4, 858 |

a. Overall denominators for condom use and HIV testing differ because eligibility criteria for answering each question differed;

b. In 2007 questions about drugs were only asked to people between 15 to 69 years;

c. Combines use of ecstasy, cocaine, or heroin

Abbreviations: m, months; SFr, Swiss Francs; yrs, years

*Table S10. Lifetime HIV testing, men and women with only opposite-sex partners. Univariable logistic regression, 2012 and 2017*

| HIV Testing | Men 2007 Unadjusted OR [a] (95% CI) | Men 2012 Unadjusted OR [a] (95% CI) | Men 2017 Unadjusted OR [a] (95% CI) | Women 2007 Unadjusted OR [a] (95% CI) | Women 2012 Unadjusted OR [a] (95% CI) | Women 2017 Unadjusted OR[a] (95% CI) |
|---|---|---|---|---|---|---|
| **Age, years** | p<0.001 | p<0.001 | p<0.001 | p<0.001 | p<0.001 | p<0.001 |
| | 0.98 (0.99, 1.00) | 0.98(0.99,1.00) | 0.98(0.99,1.00) | 0.96 (0.95, 0.96) | 0.97 (0.97, 0.97) | 0.98 (0.97, 0.98) |
| **Region** | p<0.001 | p<0.001 | p<0.001 | p<0.001 | p<0.001 | p<0.001 |
| Lake Geneva | reference | reference | reference | reference | reference | reference |
| Midland | 0.51 (0.42, 0.62) | 0.5 (0.41, 0.60) | 0.59 (0.50, 0.69) | 0.65 (0.55,0.78) | 0.55 (0.46, 0.66) | 0.54 (0.46, 0.63) |
| Northwest | 0.58 (0.46, 0.73) | 0.52 (0.42, 0.63) | 0.58 (0.48, 0.70) | 0.51 (0.42,0.64) | 0.5 (0.41, 0.61) | 0.58 (0.48, 0.69) |
| Zurich | 0.69 (0.56, 0.85) | 0.79 (0.64, 0.98) | 0.76 (0.63, 0.91) | 0.71 (0.58,0.86) | 0.63 (0.51, 0.78) | 0.67 (0.56, 0.80) |
| East | 0.39 (0.30, 0.50) | 0.46 (0.37, 0.58) | 0.52 (0.44, 0.61) | 0.4 (0.32, 0.51) | 0.47 (0.38, 0.59) | 0.41 (0.34, 0.48) |
| Central | 0.59 (0.46, 0.75) | 0.45 (0.37, 0.56) | 0.51 (0.42, 0.62) | 0.45 (0.37,0.56) | 0.44 (0.36, 0.53) | 0.37 (0.30, 0.45) |
| Ticino | 0.62 (0.47, 0.81) | 0.78 (0.61, 0.99) | 0.63 (0.51, 0.79) | 0.58 (0.46,0.73) | 0.63 (0.49, 0.79) | 0.58 (0.47, 0.71) |
| **Highest educational level** | p<0.001 | p<0.001 | p<0.001 | p<0.001 | p<0.001 | p<0.001 |
| No school/primary | reference | reference | reference | reference | reference | reference |
| Secondary | 1.55 (1.20, 2.02) | 1.69 (1.35, 2.11) | 1.49 (1.25, 1.79) | 2.18 (1.78,2.67) | 2.09 (1.69, 2.58) | 1.63 (1.37, 1.93) |
| Tertiary | 2.46 (1.89, 3.21) | 2.18 (1.74, 2.72) | 2.34 (1.95, 2.80) | 4.52 (3.59,5.70) | 4.05 (3.21, 5.11) | 3.31 (2.76, 3.96) |
| **Income, SFr** | p<0.001 | p<0.001 | p<0.001 | p<0.001 | p<0.001 | p<0.001 |
| No Income | 0.65 (0.39, 1.07) | 0.66 (0.45, 0.97) | 1.25 (0.96, 1.64) | 0.87 (0.72,1.04) | 0.91 (0.76, 1.09) | 0.96 (0.81, 1.13) |
| <4500 | reference | reference | reference | reference | reference | reference |
| 4500-6000 | 1.28 (1.07, 1.54) | 1.17 (0.98, 1.39) | 1.21 (1.05, 1.41) | 1.82 (1.51,2.19) | 1.65 (1.34, 2.03) | 1.88 (1.60, 2.21) |
| >6000 | 1.52 (1.28, 1.79) | 1.33 (1.13, 1.55) | 1.66 (1.45, 1.89) | 2.39 (1.84,3.10) | 1.89 (1.47, 2.43) | 2.34 (1.93, 2.85) |
| **Marital Status** | p<0.001 | p<0.001 | p<0.001 | p<0.001 | p<0.001 | p<0.001 |
| Single | 1.70 (1.47, 1.98) | 1.5 (1.31, 1.72) | 1.43 (1.27, 1.61) | 1.82 (1.58,2.11) | 1.5 (1.31, 1.73) | 1.65 (1.46, 1.86) |
| Married | reference | reference | reference | reference | reference | reference |
| Widowed | 0.65 (0.34, 1.22) | 1.17 (0.66, 2.08) | 1.17 (0.65, 2.10) | 0.43 (0.31,0.60) | 0.76 (0.46, 1.24) | 0.58 (0.41, 0.81) |
| Divorced | 2.91 (2.33, 3.64) | 2.27 (1.76, 2.92) | 2.41 (1.97, 2.94) | 1.98 (1.66,2.38) | 2.51 (1.99, 3.15) | 2.33 (1.96, 2.78) |
| **Age first intercourse, yrs** | p<0.001 | p<0.001 | p<0.001 | p<0.001 | p<0.001 | p<0.001 |
| | 0.90 (0.88, 0.93) | 0.91 (0.89, 0.93) | 0.97 (0.96, 0.98) | 0.88 (0.86,0.90) | 0.88 (0.85, 0.90) | 0.95 (0.93, 0.98) |
| **Sex partners last 12m** | p<0.001 | p<0.001 | p<0.001 | p<0.001 | p<0.001 | p<0.001 |
| **1** | reference | reference | reference | reference | reference | reference |
| **2-4** | 1.94 (0.85, 4.42) | 1.24 (1.00, 1.55) | 1.46 (1.20, 1.77) | 1.69 (1.31,2.19) | 1.56 (1.20, 2.02) | 2.06 (1.60, 2.67) |
| **5+** | 5.83 (2.07, 16.4) | 1.67 (1.16, 2.41) | 1.63 (1.16, 2.30) | 1.18 (0.51,2.72) | 2.12 (0.81, 5.54) | 2.71 (1.01, 7.30) |
| **Condom use last intercourse** | p=0.2 | p=0.8 | p=0.3 | p=0.3 | p= 0.8 | p= 0.051 |
| Used | reference | reference | reference | reference | reference | reference |
| Not used | 0.90 (0.76, 1.06) | 0.98 (0.84, 1.14) | 1.07 (0.94, 1.21) | 1.08 (0.92,1.28) | 1.02 (0.87, 1.20) | 1.14 (1.00, 1.29) |
| **Alcohol** | p=0.13 | p=0.8 | p<0.001 | p<0.001 | p<0.001 | p<0.001 |
| Abstinent | reference | reference | reference | reference | reference | reference |
| Daily | 1.01 (0.75, 1.35) | 0.96 (0.73, 1.25) | 0.88 (0.71, 1.10) | 0.62 (0.48,0.80) | 0.75 (0.58, 0.98) | 0.72 (0.57, 0.92) |
| Weekly | 1.21 (0.92, 1.57) | 1.02 (0.80, 1.29) | 1.27 (1.06, 1.53) | 1.27 (1.07,1.50) | 1.05 (0.88, 1.25) | 1.41 (1.23, 1.63) |
| Monthly | 1.07 (0.79, 1.43) | 1.06 (0.81, 1.39) | 1.24 (1.01, 1.53) | 1.03 (0.86,1.22) | 0.95 (0.79, 1.14) | 1.16 (1.00, 1.36) |
| **Tobacco** | p<0.001 | p<0.001 | p=0.03 | p<0.001 | p<0.001 | p<0.001 |
| Non-smoker | reference | reference | reference | reference | reference | reference |
| Casual smoker | 1.52 (1.22, 1.89) | 1.28 (1.03, 1.59) | 1.29 (1.08, 1.56) | 1.3 (1.01, 1.66) | 1.65 (1.30, 2.08) | 1.42 (1.17, 1.73) |
| Daily smoker | 1.61 (1.38, 1.88) | 1.38 (1.19, 1.59) | 1.18 (1.04, 1.34) | 1.48 (1.27,1.71) | 1.46 (1.25, 1.71) | 1.49 (1.31, 1.71) |
| **Cannabis**[b] | p<0.001 | p<0.001 | p<0.001 | p<0.001 | p<0.001 | p<0.001 |
| No use | reference | reference | reference | reference | reference | reference |

| | | | | | | |
|---|---|---|---|---|---|---|
| Any use | 2.46 (2.12, 2.86) | 2.36 (2.06, 2.71) | 2.22 (1.98, 2.50) | 3.34 (2.84,3.93) | 2.93 (2.47, 3.47) | 5.33 (3.48, 8.14) |
| **Other illicit drugs[b,c]** | p<0.001 | p<0.001 | p<0.001 | p<0.001 | p<0.001 | p<0.001 |
| No use | reference | reference | reference | reference | reference | reference |
| Any use | 4.62 (3.41, 6.27) | 2.78 (2.09, 3.69) | 2.60 (2.10, 3.23) | 6.55 (4.33,9.91) | 2.87 (2.52, 3.27) | 5.02 (3.54, 7.14) |

a. p value from Wald test;

b. In 2007 questions about drugs were only asked to people between 15 to 69 years;

c. Combines use of ecstasy, cocaine, or heroin and combines categories of use more than 12 months ago and use within last 12 months.

Abbreviations: CI, confidence interval; m, months; OR, odds ratio; SFr, Swiss Francs; yrs, years;

*Table S11. HIV testing in the last 12 months, men and women reporting only opposite-sex partners and men reporting any same-sex partner 2012, 2017.*

| **HIV testing in the last 12 months** | **Men reporting only opposite-sex partners** | | **Women reporting only opposite-sex partners** | | **Men reporting any same-sex partner** | |
|---|---|---|---|---|---|---|
| | **2012** | **2017** | **2012** | **2017** | **2012** | **2017** |
| | **Prevalence % (95% CI)** | **Prevalence % (95% CI)** | **Prevalence % (95% CI)** | **Prevalence % (95% CI)** | **Prevalence % (95% CI)** | **Prevalence % (95% CI)** |
| Total | 7 (6-8) | 7 (7-8) | 9 (8-10) | 8 (7-9) | 30 (14-38) | 39 (30-47) |
| **Age, years** | | | | | | |
| 16-24 | 14 (11-18) | 18 (15-21) | 16 (13-19) | 16 (13-19) | 35 (11-70) | 35 (17-59) |
| 25-34 | 10 (8-13) | 10 (8-12) | 18 (15-20) | 16 (13-18) | 41 (23-62) | 60 (39-79) |
| 35-44 | 7 (6-9) | 8 (6-10) | 10 (8-12) | 9 (7-11) | 32 (19-48) | 35 (19-55) |
| 45-54 | 6 (5-8) | 5 (4-6) | 4 (3-5) | 4 (3-5) | 27 (14-45) | 44 (26-63) |
| 55-64 | 3 (2-4) | 4 (3-5) | 2 (1-4) | 3 (2-4) | 26 (11-50) | 19 (9-37) |
| 65-74 | 4 (2-5) | 2 (1-3) | 1 (0-3) | 1 (1-2) | 12 (2-49) | - |
| **Number of partners** | | | | | | |
| 1 | 5 (5-6) | 5 (5-6) | 8 (7-8) | 7 (6-8) | 20 (12-30) | 31 (21-44) |
| 2-4 | 17 (13-21) | 18 (15-22) | 28 (23-34) | 31 (26-37) | 42 (22-63) | 48 (29-67) |
| 5+ | 30 (21-40) | 28 (21-36) | 41 (20-65) | 33 (16-56) | 61 (37-82) | 50 (31-69 |

*Table S12. Lifetime HIV testing, men and women with any same-sex partners. Denominators, unweighted and weighted, and number of missing values for each survey and variable, 2007, 2012, 2017*

| HIV Testing | Men | Men | Men | Women | Women | Women |
|---|---|---|---|---|---|---|
| | 2007 | 2012 | 2017 | 2007 | 2012 | 2017 |
| **Totals, N unweighted, weighted[a]** | 187, 64684 | 237, 81464 | 178,74070 | 216, 67259 | 289, 92734 | 172, 69018 |
| **Age, years** | | | | | | |
| 16-24 | 7, 3497 | 20, 5643 | 25, 8197 | 20, 10605 | 54, 15237 | 14084, 8197 |
| 25-34 | 43, 17370 | 33, 14174 | 32, 17291 | 48, 17530 | 79, 30848 | 25382, 17291 |
| 35-44 | 58, 19554 | 68, 27822 | 38, 18051 | 64, 15196 | 62, 19610 | 12090, 18051 |
| 45-54 | 36, 11628 | 59, 18085 | 36, 14631 | 56, 18019 | 61, 17425 | 12713, 14631 |
| 55-64 | 31, 9701 | 38, 11063 | 37, 13623 | 22, 4563 | 23, 6537 | 3117, 13623 |
| 65-74 | 12, 2935 | 19, 4678 | 10, 2277 | 6, 1344 | 10, 3077 | 1631, 2277 |
| **Region** | | | | | | |
| Lake Geneva | 29, 10,828 | 43, 14665 | 44, 18486 | 53, 13946 | 61, 19001 | 43, 16336 |
| Midland | 52, 17386 | 35, 15606 | 17, 8466 | 45, 11739 | 52, 17968 | 38, 18342 |
| Northwest | 17, 6329 | 43, 10864 | 19, 10322 | 34, 12394 | 56, 17660 | 16, 6521 |
| Zurich | 45, 18480 | 49, 24087 | 37, 19055 | 44, 17850 | 40, 21183 | 26, 14769 |
| East | 18, 5623 | 27, 9230 | 27, 10121 | 17, 5344 | 29, 7467 | 20, 6475 |
| Central | 14, 3722 | 34, 5939 | 26, 5903 | 19, 5549 | 36, 6356 | 20, 4442 |
| Ticino | 12, 2317 | 6, 1073 | 8, 1718 | 4, 437 | 15, 3100 | 9, 2133 |
| **Highest educational level** | | | | | | |
| No school/ primary | 8, 3259 | 19, 4915 | 15, 4467 | 10, 2791 | 27, 8459 | 17, 5036 |
| Secondary | 98, 30528 | 106, 37877 | 80, 31484 | 131, 43428 | 152, 51662 | 83, 33636 |
| Tertiary | 81, 30897 | 112, 38672 | 83, 38119 | 75, 21040 | 110, 32612 | 72, 30346 |
| Missing | 0, 0 | 0, 0 | 0, 0 | 0, 0 | 0, 0 | 0, 0 |
| **Income, SFr** | | | | | | |
| No Income | 5, 2205 | 9, 2488 | 12, 3984 | 14, 5575 | 19, 6716 | 8, 2581 |
| <4500 | 66, 24662 | 64, 20121 | 62, 26067 | 121, 38545 | 178, 57005 | 106, 44156 |
| 4500-6000 | 51, 16664 | 49, 16453 | 42, 17768 | 42, 12900 | 37, 11380 | 31, 10373 |
| >6000 | 51, 17320 | 732, 31517 | 50, 20141 | 24, 5869 | 29, 9141 | 22, 9965 |
| Missing | 14, 3833 | 32, 10885 | 12, 6110 | 15, 4370 | 26, 8492 | 5, 1943 |
| **Marital status** | | | | | | |
| Single | 119, 41500 | 118, 45409 | 120, 52194 | 117, 35396 | 147, 48911 | 111, 47453 |
| Married | 41, 14123 | 96, 26288 | 18, 5042 | 47, 18229 | 99, 27702 | 22, 7883 |
| Widowed | 0, 0 | 1, 178 | 0, 0 | 4, 839 | 5, 1679 | 2, 428 |
| Divorced | 26, 7948 | 22, 9589 | 13, 5267 | 48, 12795 | 37, 13873 | 14, 5518 |
| Missing | 1, 1113 | 0, 0 | 27, 11566 | 0, 0 | 1, 569 | 23, 7735 |
| **Sex partners last 12m** | | | | | | |
| 1 | 78, 28188 | 158,51901 | 86,34681 | 136, 45498 | 3367, 75583 | 106, 44848 |
| 2-4 | 62, 21451 | 44,17482 | 40,19451 | 36, 10840 | 253, 13385 | 41, 16743 |
| 5+ | 26, 10147 | 30,10335 | 37,14688 | 12, 3257 | 23, 3767 | 7, 3005 |
| Missing | 8, 4889 | 5, 1747 | 15, 5250 | 32, 7664 | 0, 0 | 18, 4422 |
| **Condom use last intercourse** | | | | | | |
| Used | 95, 35009 | 150,53233 | 91,34347 | 144, 46564 | 213, 66891 | 129, 51731 |
| Not used | 80, 26642 | 86,27692 | 87,39903 | 51, 14040 | 76, 25843 | 43, 17287 |
| Missing | 8, 3033 | 1, 539 | 0, 0 | 21, 6656 | 0, 0 | 0, 0 |
| **Alcohol** | | | | | | |
| No use | 17, 4041 | 28, 8021 | 12, 4354 | 20, 7383 | 32, 10546 | 225, 72098 |
| Daily | 37, 14060 | 36, 12419 | 28, 9542 | 23, 6067 | 24, 7291 | 64, 20636 |
| Weekly | 96, 31944 | 132, 46296 | 99, 44739 | 114, 35551 | 155, 48066 | 225, 72098 |
| Monthly | 37, 14638 | 41, 14729 | 39, 15436 | 58, 18006 | 78, 26831 | 64, 20636 |
| Missing | 0, 0 | 0, 0 | 0, 0 | 1, | 0, 0 | 0, 0 |
| **Tobacco** | | | | | | |
| Non-smoker | 102, 37373 | 137, 46754 | 103, 43499 | 102,37373 | 139, 47724 | 84, 34367 |

| | | | | | | |
|---|---|---|---|---|---|---|
| Casual smoker | 25, 7274 | 30, 11727 | 18, 5952 | 25,7274 | 50, 16277 | 19, 7859 |
| Daily smoker | 60, 20037 | 70, 22983 | 57, 24620 | 60,20037 | 100, 28733 | 69, 26792 |
| Missing | 0, 0 | 0, 0 | 0, 0 | 0, 0 | 0, 0 | 0, 0 |
| **Cannabis**[b] | | | | | | |
| No use | 90, 26874 | 125, 40575 | 91, 37982 | 91, 27166 | 86, 28336 | 74, 28862 |
| Any use | 90, 35547 | 110, 39979 | 85, 34967 | 89, 27750 | 203, 64398 | 96, 39422 |
| Missing | 7, 2264 | 2, 910 | 2, 1121 | 3, 1547 | 0, 0 | 2,734 |
| **Other illicit drugs**[b, c] | | | | | | |
| No use | 138, 46,062 | 178, 57781 | 144, 60100 | 138,52314 | 225, 72,098 | 136, 53648 |
| Any use | 43, 16,637 | 54, 22233 | 34, 13970 | 43,14945 | 64, 20,636 | 36, 15370 |
| Missing | 6, 19860 | 5, 1543 | 0, 0 | 0, 0 | 0, 0 | 0, 0 |

a. Overall denominators for condom use and HIV testing differ because eligibility criteria for answering each question differed;

b. In 2007 questions about drugs were only asked to people between 15 to 69 years;

c. Combines use of ecstasy, cocaine, or heroin

Abbreviations: m, months; SFr, Swiss Francs; yrs, years

*Table S13. Lifetime HIV testing, men reporting any same-sex partner. Associations with sociodemographic factors, sexual behaviours and substance use from univariable logistic regression, 2007, 2012, 2017*

| **HIV testing** | **2007** | **2012** | **2017** |
|---|---|---|---|
| | **Unadjusted OR (95% CI)** [a] | **Unadjusted OR (95% CI)** [a] | **Unadjusted OR (95% CI)** [a] |
| **Age** | p=0.072 | p=0.3 | p=0.9 |
| **Per year increase** | 0.97 (0.94, 1.00) | 0.98 (0.93, 1.02) | 1.00 (0.97, 1.04) |
| **Region** | p=0.2 | p=0.049 | p<0.001 |
| Lake Geneva | reference | reference | reference |
| Midland | 0.71 (0.21, 2.37) | 0.91 (0.28, 3.03) | 0.66 (0.12, 3.78) |
| Northwest | 1.81 (0.35, 9.48) | 0.46 (0.12, 1.75) | 0.47 (0.11, 1.91) |
| Zurich | 1.16 (0.33, 4.11) | 2.93 (0.75, 11.5) | 1.82 (0.47, 7.07) |
| East | 0.24 (0.05, 1.03) | 0.3 (0.08, 1.07) | 0.57 (0.15, 2.15) |
| Central | 1.8 (0.23, 14.2) | 0.46 (0.13, 1.62) | 0.39 (0.11, 1.41) |
| Ticino | 2.42 (0.37, 15.7) | 0.40 (0.06, 2.83) | - |
| **Education level** | p=0.8 | p=0.01 | p=0.10 |
| No school/primary | reference | reference | reference |
| Secondary | 0.86 (0.12, 6.06) | 0.63 (0.16, 2.41) | 0.63 (0.16, 2.41) |
| Tertiary | 1.11 (0.16, 7.73) | 1.84 (0.43, 7.89) | 1.84 (0.43, 7.89) |
| **Income, SFr** | p=0.6 | p=0.14 | p=0.011 |
| No Income | 0.69 (0.08, 6.37) | 1.15 (0.22, 5.94) | 0.66 (0.16, 2.64) |
| <4500 | reference | reference | reference |
| 4500-6000 | 0.79 (0.28, 2.25) | 2.91 (1.02, 8.31) | 3.85 (1.20, 12.4) |
| >6000 | 0.50 (0.18, 1.40) | 2.48 (0.95, 6.47) | 4.2 (1.29, 13.7) |
| **Marital Status** | p=0.001 | p<0.001 | p=0.8 |
| Single | 4.51 (1.84, 11.1) | 2.15 (0.96, 4.81) | 0.95 (0.26, 3.42) |
| Married | reference | reference | reference |
| Widowed | - | - | - |
| Divorced | 7.34 (1.88, 28.7) | 0.83 (0.19, 3.61) | 1.58 (0.26, 9.49) |
| **Age first sexual intercourse, years** | p= 0.038 | p= 0.9 | p= 0.5 |
| **Median (IQR)** | 0.87 (0.76, 0.99) | 0.99 (0.88, 1.11) | 0.97 (0.89, 1.06) |
| **Sex partners 12m** | p<0.001 | p=0.6 | p=0.3 |
| **1** | reference | reference | reference |
| **2-4** | 1.26 (0.50, 3.18) | 1.40 (0.53, 3.70) | 2.29 (0.75, 6.96) |
| **5+** | 27.1 (4.99, 147) | 2.43 (0.38, 15.7) | 0.98 (0.29, 3.39) |
| **Condom use last intercourse** | p=0.072 | p=0.2 | p=0.8 |
| Used | reference | reference | reference |
| Not used | 2.18 (0.93, 5.12) | 1.95 (0.78, 4.89) | 0.87 (0.37, 2.08) |
| **Alcohol** | p=0.061 | p=0.090 | p=0.11 |
| No use | reference | reference | reference |
| Daily | 0.09 (0.01, 0.56) | 1.37 (0.31, 6.13) | 1.27 (0.26, 6.18) |
| Weekly | 0.19 (0.03, 1.11) | 3.16 (1.04, 9.63) | 2.66 (0.63, 11.2) |
| Monthly | 0.13 (0.02, 0.88) | 3.52 (0.93, 13.3) | 5.64 (1.04, 30.7) |
| **Tobacco** | p=0.3 | p=0.2 | p=0.68 |
| Non-smoker | reference | reference | reference |
| Casual smoker | 0.69 (0.23, 2.12) | 0.55 (0.18, 1.72) | 1.03 (0.29, 3.57) |
| Daily smoker | 1.80 (0.67, 4.84) | 1.71 (0.71, 4.09) | 3.24 (1.16, 9.08) |
| **Cannabis** | p=0.023 | p=0.010 | p=0.9 |
| No use | reference | reference | reference |
| Any use | 3.22 (1.36, 7.63) | 3.86 (1.37, 10.8) | 0.94 (0.39, 2.26) |
| **Other illicit drugs**[b] | p=0.019 | p= 0.002 | p=0.7 |
| No use | reference | reference | reference |
| Any use | 4.86 (1.28, 18.4) | 4.36 (1.67, 11.4) | 1.35 (0.32, 5.70) |

a. p value from Wald test;

b. In 2007 questions about drugs were only asked to people between 15 to 69 years;

c. Combines use of ecstasy, cocaine, or heroin and combines categories of use more than 12 months ago and use within last 12 months.

Abbreviations: CI, confidence interval; m, months; OR, odds ratio; SFr, Swiss Francs; yrs, years;

*Table S14. Lifetime HIV testing, women reporting any same-sex partner. Associations with sociodemographic factors, sexual behaviours and substance use from univariable logistic regression, 2007, 2012, 2017*

| HIV testing | 2007 | | 2012 | | 2017 | | |
|---|---|---|---|---|---|---|---|
| | Prevalence (95% CI) | Unadjusted OR (95% CI) | Prevalence (95 CI%) | Unadjusted OR (95% CI) | Prevalence (95 CI%) | Unadjusted OR (95% CI) | Adjusted OR (95% CI) [a] |
| **Year** | | | | | | | p=0.5 |
| 2007 | | | | | | | reference |
| 2012 | | | | | | | 1.35 (0.82, 2.23) |
| 2017 | | | | | | | 1.31 (0.74, 2.30) |
| **Age** | | p=0.020 | | p=0.7 | | p=0.8 | p=0.9 |
| Median age (years) (IQR) | 36 (26-45) | 0.96 (0.93, 0.99) | 34(29-45) | 0.99 (0.97, 1.02) | 33 (26-44) | 1.01 (0.97, 1.04) | 1 (0.98, 1.02) |
| 16-24 | 70 (38-90) | - | 48 (32-65) | - | 59 (37-77) | - | - |
| 25-34 | 70 (54-83) | - | 82 (69-90) | - | 67 (51-80) | - | - |
| 35-44 | 71 (55-83) | - | 79 (64-89) | - | 88 (68-96) | - | - |
| 45-54 | 51 (33-68) | - | 68 (52-81) | - | 75 (55-88) | - | - |
| 55-64 | 25 (09-54) | - | 63 (31-87) | - | 45 (05-93) | - | - |
| **Region** | | | | p=0.5 | | p=0.3 | |
| Lake Geneva | 60 (43-76) | reference | 68 (52-80) | reference | 71 (53-84) | reference | .. |
| Midland | 64 (45-80) | 1.14 (0.41, 3.21) | 77 (59-88) | 1.58 (0.57, 4.41) | 64 (46-80) | 0.74 (0.26, 2.12) | .. |
| Northwest | 63 (40-82) | 1.1 (0.35, 3.41) | 57 (41-72) | 0.64 (0.26, 1.58) | 56 (28-81) | 0.53 (0.14, 2.01) | .. |
| Zurich | 54 (35-72) | 0.77 (0.28, 2.09) | 71 (52-85) | 1.18 (0.42, 3.31) | 80 (55-93) | 1.69 (0.43, 6.57) | .. |
| East | 73 (38-94) | 1.73 (0.37, 8.12) | 79 (52-93) | 1.79 (0.46, 6.94) | 62 (29-87) | 0.68 (0.15, 2.99) | .. |
| Central | 67 (29-93) | 1.31 (0.26, 6.57) | 78 (59-89) | 1.66 (0.57, 4.84) | 81 (55-94) | 1.74 (0.45, 6.77) | .. |
| Ticino | 43 (0-100) | 0.48 (0.04, 5.85) | 75 (41-93) | 1.43 (0.32, 6.37) | 37(10-76) | 0.24 (0.05, 1.22) | .. |
| **Education level** | | p=0.2 | | p=0.5 | | =0.3 | |
| No school/primary | 30 (04-84) | reference | 56 (31-78) | reference | 68 (34-90) | reference | .. |
| Secondary | 65 (54-75) | 4.28 (0.87, 21.0) | 69 (59-78) | 1.78 (0.64, 4.93) | 67 (53-78) | 0.92 (0.26, 3.28) | .. |
| Tertiary | 57 (43-70) | 3.03 (0.59, 15.5) | 76 (64-84) | 2.46 (0.85, 7.07) | 71 (57-81) | 1.11 (0.30, 4.07) | .. |
| **Income, SFr** | | p=0.5 | | p=0.7 | | p=0.3 | |
| No Income | 80 (39-96) | 2.38 (0.47, 12.1) | 86 (48-98) | 3.48 (0.68, 17.8) | 60 (13-94) | 0.9 (0.17, 4.83) | .. |
| <4500 | 62 (50-73) | reference | 64 (55-72) | reference | 63 (51-73) | reference | .. |
| 4500-6000 | 56 (35-75) | 0.78 (0.30, 2.04) | 64 (43-80) | 1.00 (0.41, 2.43) | 67 (44-84) | 1.19 (0.44, 3.22) | .. |
| >6000 | 51 (26-76) | 0.63 (0.20, 1.94) | 91 (75-97) | 5.69 (1.69, 19.2) | 93 (75-98) | 7.84 (1.88, 32.8) | .. |
| **Marital Status** | | p=0.7 | | p=0.2 | | p=0.9 | |
| Single | 63 (51-74) | 1.05 (0.46, 2.41) | 73 (63-81) | 1.48 (0.73, 2.96) | 71 (60-80) | 2.57 (0.87, 7.54) | .. |
| Married | 62 (45-76) | reference | 64 (51-76) | reference | 49 (24-74) | reference | .. |
| Widowed | 29 (0-80) | 0.26 (0.02, 3.21) | 76 (35-100) | 1.74 (0.17, 17.5) | 45 (0-100) | 0.86 (0.04, 16.6) | .. |
| Divorced | 56 (35-75) | 0.78 (0.27, 2.26) | 71 (51-86) | 1.26 (0.43, 3.64) | 63 (21-92) | 1.81 (0.35, 9.35) | .. |
| **Age first sexual intercourse, years** | | p=0.4 | | p=0.014 | | p<0.001 | |
| **Median (IQR)**[b] | 17 (15-18) | 0.95 (0.84, 1.06) | 16 (15-18) | 0.86 (0.76, 0.97) | 16 (15-18) | 0.76 (0.64, 0.89) | .. |
| **Sex partners 12m** | | p=0.3 | | p=0.3 | | p<0.001 | p=0.026 |
| **1** | 61 (51-71) | reference | 68 (60-75) | reference | 66 (54-76) | reference | reference |
| **2-4** | 70 (48-86) | 1.47 (0.55, 3.95) | 80 (63-91) | 1.92 (0.77, 4.80) | 79 (60-91) | 2 (0.73, 5.52) | 1.79 (1.01, 3.17) |
| **5+** | 87 (31-99) | 4.32 (0.50, 37.3) | 83 (27-99) | 2.37 (0.29, 19.5) | 100 | - | 4.65 (1.02, 21.1) |
| **Condom use last intercourse** | | p=0.4 | | p=0.9 | | p=0.8 | .. |
| Used | 61 (50-70) | reference | 70 (62-77) | reference | 68 (58-77) | reference | .. |
| Not used | 70 (52-83) | 1.50 (0.63, 3.58) | 70 (56-81) | 0.99 (0.49, 2.03) | 70 (51-84) | 1.12 (0.46, 2.72) | .. |
| **Alcohol** | | p >0.9 | | p=0.4 | | p<0.001 | .. |
| No use | 66 (36-88) | reference | 55 (32-76) | reference | 95 (83-99) | reference | .. |
| Daily | 61 (33-83) | 0.79 (0.16, 3.77) | 64 (35-85) | 1.44 (0.35, 5.89) | 46 (11-87) | 0.05 (0.01, 0.31) | .. |
| Weekly | 60 (48-71) | 0.75 (0.22, 2.60) | 72 (63-80) | 2.13 (0.78, 5.80) | 71 (60-81) | 0.13 (0.03, 0.52) | .. |
| Monthly | 62 (44-77) | 0.82 (0.22, 3.12) | 74 (60-84) | 2.30 (0.78, 6.81) | 54 (36-71) | 0.06 (0.01, 0.27) | .. |
| **Tobacco** | | p=0.4 | | p=0.6 | | p= 0.5 | .. |
| Non-smoker | 60 (47-71) | reference | 68 (58-77) | reference | 67 (54-78) | reference | .. |
| Casual smoker | 48 (23-73) | 0.61 (0.20, 1.86) | 67 (49-81) | 0.41, 2.21 | 80 (50-94) | 1.96 (0.58, 6.63) | .. |
| Daily smoker | 66 (51-78) | 1.27 (0.58, 2.76) | 75 (63-85) | 0.68, 2.97 | 68 (53-80) | 1.04 (0.46, 2.35) | .. |
| **Cannabis**[c] | | p=0.11 | | p=0.001 | | p<0.001 | .. |
| No use | 50 (37-63) | reference | 53 (40-66) | reference | 51 (37-64) | reference | .. |
| Any use | 70 (57-80) | 2.31 (1.06, 5.06) | 78 (69-84) | 3.04 (1.54, 5.97) | 82 (71-90) | 4.46 (1.92, 10.4) | .. |
| **Other illicit drugs**[d] | | p=0.3 | | p=0.13 | | p=0.2 | .. |
| No use | 59 (49-68) | reference | 67 (59-74) | reference | 65 (55-74) | reference | .. |
| Any use | 69 (50-83) | 1.55 (0.65, 3.69) | 81 (63-92) | 2.09 (0.79, 5.53) | 80 (58-93) | 2.21 (0.71, 6.89) | .. |

a. Multivariable model includes age, and number of sex partners; p value from Wald test;

b. Median age of the proportion tested for HIV;

c. In 2007 questions about drugs were only asked to people between 15 to 69 years;

d. Combines use of ecstasy, cocaine, or heroin and combines categories of use more than 12 months ago and use within last 12 months.

Abbreviations: CI, confidence interval; IQR, interquartile range; m, months; OR, odds ratio; SFr, Swiss Francs; yrs, years; .., variable not included in multivariable model.
