## Supplementary material for "Condom use and HIV testing among adults in Switzerland: repeated national cross-sectional surveys 2007, 2012, and 2017": STROBE checklist

STROBE Statement—Checklist of items that should be included in reports of ***cross-sectional studies***

| | Item No | Recommendation | Location where item is reported |
|---|---|---|---|
| **Title and abstract** | 1 | (*a*) Indicate the study's design with a commonly used term in the title or the abstract | Page 1 |
| | | (*b*) Provide in the abstract an informative and balanced summary of what was done and what was found | Page 2 |
| **Introduction** | | | |
| Background/rationale | 2 | Explain the scientific background and rationale for the investigation being reported | Page 4 |
| Objectives | 3 | State specific objectives, including any prespecified hypotheses | Page 4-5 |
| **Methods** | | | |
| Study design | 4 | Present key elements of study design early in the paper | Page 5 |
| Setting | 5 | Describe the setting, locations, and relevant dates, including periods of recruitment, exposure, follow-up, and data collection | Page 5 |
| Participants | 6 | (*a*) Give the eligibility criteria, and the sources and methods of selection of participants | Page 5<br>Table S1 |
| Variables | 7 | Clearly define all outcomes, exposures, predictors, potential confounders, and effect modifiers. Give diagnostic criteria, if applicable | Page 6 |
| Data sources/ measurement | 8* | For each variable of interest, give sources of data and details of methods of assessment (measurement). Describe comparability of assessment methods if there is more than one group | Page 6 |
| Bias | 9 | Describe any efforts to address potential sources of bias | Page 5 |
| Study size | 10 | Explain how the study size was arrived at | Page 5 |
| Quantitative variables | 11 | Explain how quantitative variables were handled in the analyses. If applicable, describe which groupings were chosen and why | Page 6-7 |
| Statistical methods | 12 | (*a*) Describe all statistical methods, including those used to control for confounding | Page 6-7 |
| | | (*b*) Describe any methods used to examine subgroups and interactions | Page 6-7 |
| | | (*c*) Explain how missing data were addressed | Page 7 |
| | | (*d*) If applicable, describe analytical methods taking account of sampling strategy | Page 6 |

| | | | |
|---|---|---|---|
| | | (*e*) Describe any sensitivity analyses | Not Applicable |
| **Results** | | | |
| Participants | 13* | (a) Report numbers of individuals at each stage of study—eg numbers potentially eligible, examined for eligibility, confirmed eligible, included in the study, completing follow-up, and analysed | Page 7-8 |
| | | (b) Give reasons for non-participation at each stage | Page 8<br>Table S1 |
| | | (c) Consider use of a flow diagram | Page 8 |
| Descriptive data | 14* | (a) Give characteristics of study participants (eg demographic, clinical, social) and information on exposures and potential confounders | Page 8-19 |
| | | (b) Indicate number of participants with missing data for each variable of interest | Supp tables |
| Outcome data | 15* | Report numbers of outcome events or summary measures | |
| Main results | 16 | (*a*) Give unadjusted estimates and, if applicable, confounder-adjusted estimates and their precision (eg, 95% confidence interval). Make clear which confounders were adjusted for and why they were included | Table 2-5<br>Supp tables |
| | | (*b*) Report category boundaries when continuous variables were categorized | Table 2-5 |
| | | (*c*) If relevant, consider translating estimates of relative risk into absolute risk for a meaningful time period | Not applicable |
| Other analyses | 17 | Report other analyses done—eg analyses of subgroups and interactions, and sensitivity analyses | Not applicable |
| **Discussion** | | | |
| Key results | 18 | Summarise key results with reference to study objectives | Page 19-20 |
| Limitations | 19 | Discuss limitations of the study, taking into account sources of potential bias or imprecision. Discuss both direction and magnitude of any potential bias | Page 20 |
| Interpretation | 20 | Give a cautious overall interpretation of results considering objectives, limitations, multiplicity of analyses, results from similar studies, and other relevant evidence | Page 21 |
| Generalisability | 21 | Discuss the generalisability (external validity) of the study results | Page 22-23 |
| **Other information** | | | |
| Funding | 22 | Give the source of funding and the role of the funders for the present study and, if applicable, for the original study on which the present article is based | Page 24 |
